# Divergent Neuroimmune Signatures in the Cerebrospinal Fluid Predict Differential Gender-Specific Survival Among Patients With HIV-Associated Cryptococcal Meningitis

**DOI:** 10.1101/2023.08.09.23293903

**Authors:** Samuel Okurut, David R Boulware, Elizabeth Okafor, Joshua Rhein, Henry Kajumbula, Bernard Bagaya, Freddie Bwanga, Joseph O Olobo, Yukari C Manabe, David B Meya, Edward N Janoff, ASTRO trial team

## Abstract

Survival among people with HIV-associated cryptococcal meningitis (CM) remains low, exceptionally among women with the increased threat of death on current optimal use of antifungal drugs. *Cryptococcus* dissemination into the central nervous system (CNS) prompts a neuroimmune reaction to activate pathogen concomitant factors. However, no consistent diagnostic or prognostic immune-mediated signature is reported to underpin the risk of death or mechanism to improve treatment or survival. We theorized that the distinct neuroimmune cytokine or chemokine signatures in the cerebrospinal fluid (CSF), distinguish survivors from people who died on antifungal treatment, who may benefit from tailored therapy. We considered the baseline clinical disease features, cryptococcal microbiologic factors, and CSF neuroimmune modulated signatures among 419 consenting adults by gender (biological sex assigned at birth) (168 females and 251 males) by 18 weeks of survival on antifungal management. Survival at 18 weeks was inferior among females than males (47% vs. 59%; hazard ratio HR=1.4, 95% CI: 1.0 to 1.9, and p=0.023). Unsupervised principal component analysis (PCA) demonstrated the divergent neuroimmune signatures by gender, survival, and intragender-specific survival. Overall, females displayed lower levels of PD-L1, IL-1RA, and IL-15 than males (all p≤0.028). Female survivors compared with those who died, expressed significant fold elevations in levels of CSF (CCL11 - myeloid and CXCL10 - lymphoid chemokine (in both p=0.001), and CSF Th1, Th2, and Th17 cytokines. In contrast, male survivors expressed distinctly lower levels of CSF IL-15 and IL-8 compared with those who died. Survivors of either gender demonstrated a significant increase in the levels of immune regulatory element, IL-10. In the finale, we classified divergent neuroimmune key signatures in CSF by gender, survival, and intragender-specific survival among people with HIV-associated cryptococcal meningitis. These intragender-specific survival associated-neuroimmune signatures, suggests the discrete role of gender immune regulating mechanisms as the possible targets for interventions to advance therapy to improve survival among people with HIV-associated cryptococcal meningitis.

## 1 INTRODUCTION

Co-infection with the fungus *Cryptococcus neoformans* remains an important contributor to death among people with advanced HIV/AIDS worldwide, reported with the existing ideal use of antifungal medications ^1,2^. The reported global HIV-associated cryptococcal meningitis (CM) survival rates vary by location based on existing HIV prevalence rates. In Europe and North America, with the low prevalence of people with HIV, both the incidence and mortality rates of CM are lowest ^3^, in contrast, to low and middle-income countries, especially in Africa with the highest prevalence of people with HIV ^3–5^. In countries with high frequency of people with HIV in Africa, like Uganda, Botswana, and South Africa, the 10-week survival rates can fall below 50% ^3,5–8^ reported among closely monitored research settings, with deaths occurring within days to weeks, and sometimes, up to months after diagnosis ^2,9,10^. The high mortality among closely monitored patients with CM may be worrisome, for patients undertaking treatment in public health settings where specialized diagnosis and close treatment monitoring may be a challenge. These observations rank highly the importance of early immune response as possible intervention to control fungal infection in supporting better survival ^10^.

The Casadevall and Pirofski damage response framework paradigm points to the role of the pathological processes in influencing host response mechanisms to shape the infection and disease outcome in sequelae ^11,12^. That the replicating pathogen eliciting unregulated immune activated mechanism (role of immune regulation), mediate pathways that lead to host self-directed damage responses that lead to poor outcome. And that replicating pathogen in a state with immune inhibition or in the absence of the pathogen-specific induced response (case of severe immune suppression), that pathogen-associated virulence factors lead to host damage responses that lead to poor outcomes. Suggesting that improved outcome is achievable in a combined therapy involving an effective pathogen-specific target therapy (e.g., antifungals) combined with immune-based treatment to modulate immune homeostasis.

In the immunopathogenesis observations, cryptococcal dissemination into the central nervous system (CNS) tissues and CSF across the blood-brain barrier ^13–15^ leads to activation of CNS resident neuroimmune cells (astrocytes, microglial cells, local macrophages, dendritic cells, and lymphocytes). In the process, locally activated immune cells produce activating chemoattractant proteins (e.g., CCL11/Eotaxin, CXCL10/Interferon inducible protein 10 (IP-10) ^16,17^ and other inflammatory mediators (e.g., IL-15, IL-8/CXCL8) ^18^. During cryptococcal infection, CNS infiltrating immunocytes, especially T and B cells ^19,20^, may extravasate blood vessels and lymphatic channels to induce neuroimmune activation, inflammation, and meningoencephalitis. Intracerebral and intrathecal activated cytokine and chemokine production influence neuroimmune milieu-mediated responses, that shape fungal clearance, ensuing immunopathological processes, clinical phenotype, and death outcome^21–25^.

Among studies reporting cases by gender, the majority of cryptococcal meningitis is diagnosed among males ^2,7,9,19,26–44.^ Of note, few studies to date since widespread ART rollout specifically address selective survival by gender (only five including the current study) ^7^. Only recent data, from 2021 reveal that the proportion of females who succumb to fungal disease was higher than in males even in the presence of optimized antifungal and antiretroviral therapy ^9^. Since widespread ART rollout, no such differences in gender-specific survival were reported ^26,44^. To date, no clear immunopathogenic mechanisms have been proposed to explain the divergence in survival by gender despite access to current optimal antifungal treatment and ART.

In CM, males show increased expression of innate chemokines and cytokines in the CSF associated with increased trafficking of innate lymphoid and myeloid cells compared with females, ^7^. However, these differences were not evaluated in relation to gender-specific survival. Furthermore, the differences in the host responses to the vaccine induced antigens have been reported by gender in both humans and animals ^45,46^, showing elevation in cytokine production, endocrine, and metabolic parameters in females compared with males. Nevertheless, in cryptococcal meningitis, the cryptococcal fungal burden, white cell counts, and protein in CSF, and the white cells, and CD4^+^ T cell numbers in blood were similar by gender^7^. The cryptococcal host immune evasion mediating factors, e.g., *Cryptococcus* fungal vomocytosis and the cryptococcal intracellular fungal proliferation in the cryptococcal infected macrophages, were similar by gender ^47^.

We evaluated CSF-specific soluble cytokine, chemokine, and immunoregulatory responses to cryptococcal meningitis between females and males overall, and the differences in these neuroimmune responses in relation to survival by gender (Biological sex assigned at birth). We hypothesized that CSF neuroimmune cytokine and chemokine signatures would differ between females and males and across survival. In the CSF at baseline, we characterized representative Th1, Th2, Th17, Tfh cytokine, and innate myeloid–neutrophils activation regulating cytokines, and immune checkpoint markers (PD-L1) among people who survived or died during the 18 weeks of follow-up. We identified discrete differences in patterns of neuroimmune mediators by gender and by intragender-specific survival at the site of infection in the CSF.

## 2 MATERIALS AND METHODS

### 2.1 Participants, Sites, and Setting

We enrolled 419 participants from a cohort of adults with CM enrolled to receive meningitis treatment in the Adjunctive Sertraline for the Treatment of HIV-Associated Cryptococcal Meningitis trial (ClinicalTrials.gov: NCT01802385). The trial was conducted between 2015 – 2017 at the Infectious Diseases Institute (Administrative site) and at Mulago and Kiruddu National Referral Hospitals (patient catchment sites) in Kampala, Uganda ^2,48^. The participants were selected based on gender and available survival-specific data for cross-sectional analyses with reference to 18 weeks of survival after cryptococcal meningitis diagnosis. The majority (94%; 168 females and 251 males) had complete demographic, clinical, and CSF datasets. Participants or their surrogates gave informed written and signed consent under protocols approved by the institutional review board of Makerere University and the University of Minnesota Medical School. Enrolled in the study were people ≥18 years of age, with a confirmed diagnosis of HIV-associated cryptococcal meningitis co-infection as previously described ^49,50^. Only participants whose survival status was known at censoring or at study termination at 18 weeks of follow-up (379 of 419; 90.5%) were included in the survival sensitivity analyses. Of note, parent cohort survival did not differ statistically between either sertraline randomized participants or those on standard therapy alone ^2^.

### 2.2 Specimen Preparation

CSF was drawn from lumbar punctures at diagnosis of CM prior to antifungal therapy initiation. The CSF specimens were spun to pellet out cells. The CSF supernatant was stored in a −80°C freezer prior to thawing for testing using Luminex. At enrolment, all participants in the current study were antifungal treatment naïve.

### 2.3 Luminex Cytokine and Chemokine Immunophenotyping

A representative sample of cytokines, chemokines, and checkpoint regulators was measured in CSF diluted in a 1:2 ratio based on the R&D Human XL Cytokines Discovery Premixed Kit platform as per the manufacturer’s recommendations (R&D, Minneapolis, MN). The T helper 1 (Th1) cytokines were tumor necrosis factor-alpha (TNF-α), interferon-gamma (IFN-γ), interleukin 2 (IL-2), IL-12p70, with soluble CD40-Ligand/TNFSF5. The Th2 cytokines were IL-4 and IL-13. T follicular helper cytokines were IL-6 and IL-10. The Th17 cytokines included was IL-17A. Cytokines derived from innate lymphoid and myeloid cells were IL-15, IL-8, IL-1 RA/IL-1 F3. The inflammatory mediating chemokines primarily derived from microglial and astrocytes mediating neuroinflammation in cryptococcal meningitis were CXCL10 lymphoid cells mediating chemoattraction ^51,52^ and CCL11 mediating myeloid cells (Eosinophils) chemoattraction ^53,54^. The IL-8 mediating neutrophils activation and chemoattraction ^55^. The immune checkpoint molecules were programmed death ligand 1 (PD-L1/B7-H1) ^56^ and immune regulatory cytokine IL-10 ^57^.

### 2.4 Statistical Analysis

Data were analyzed using GraphPad Prism version 9.3.0, GraphPad Software, LLC for Macintosh (San Diego, California, USA). The databases were compiled using Microsoft Excel. The data variability was visualized using unsupervised Principal Component Analysis, (PCA) using Eigenvector covariates projection on biplots as described elsewhere ^58–61^. Further interrogation of the individual or independent principal component clustering and variability, factor differences, and interactions or associations of independent predictor variables by gender and by survival were performed using univariate and multivariate analyses.

Univariate analytic methods comprised pairwise comparisons using Mann-Whitney non-parametric U test that compared arithmetic medians and unpaired parametric t-test that compared arithmetic means. In this context of non-normally distributed population variables, statistical differences were reported based on the difference in the sample medians with interquartile ranges (IQR). The univariant difference in the survival outcome or survival risk was determined using Kaplan-Meier/Log-rank test (Mantel-Cox Chi-Square test) or Mann-Whitney U-test. The difference in binary outcome was determined using Fisher’s exact Chi-square test or Mann-Whitney U-test.

In multivariate models, data were Log_2_ transformed to normalize the variable prior to the statistical interrogation using multivariate factor analysis and or survival-adjusted logistic regression least square models that measure the risk of likelihood (proportional hazard ratios). In all models, missingness was not imputed. In this context, statistical significance in both the univariate and the multivariate models were based on difference among variables in the original available dataset among participants. The statistical level of the significantly different covariates was reported at a p-value <0.05 and at a 95% level of confidence. The rigor of our interrogations included a large sample size (N=419), controlled comparative covariables at nearly a 1:1 ratio, and minimal missingness with approximately 94% data completeness in gender analyses and approximately 90% data completeness in the 18 weeks of survival analyses.

## 3 RESULTS

### 3.1 Baseline Demographics, by Gender

Consistent with published reports (**Figure 1**), among 419 participants studied with HIV-associated CM, the majority were males, who were older and heavier than females (**Table 1**). About half of these participants were on ART for a median of 1.6 months (IQR; 0-22 months). Neurologic abnormalities predominated among clinical signs and symptoms, with almost all describing headache for one to four weeks, a third reporting changes in mental status, and a half with abnormal Glasgow Coma Score (GCS<15), with each variable reported at comparable frequencies in males and females.

**Table 1.**
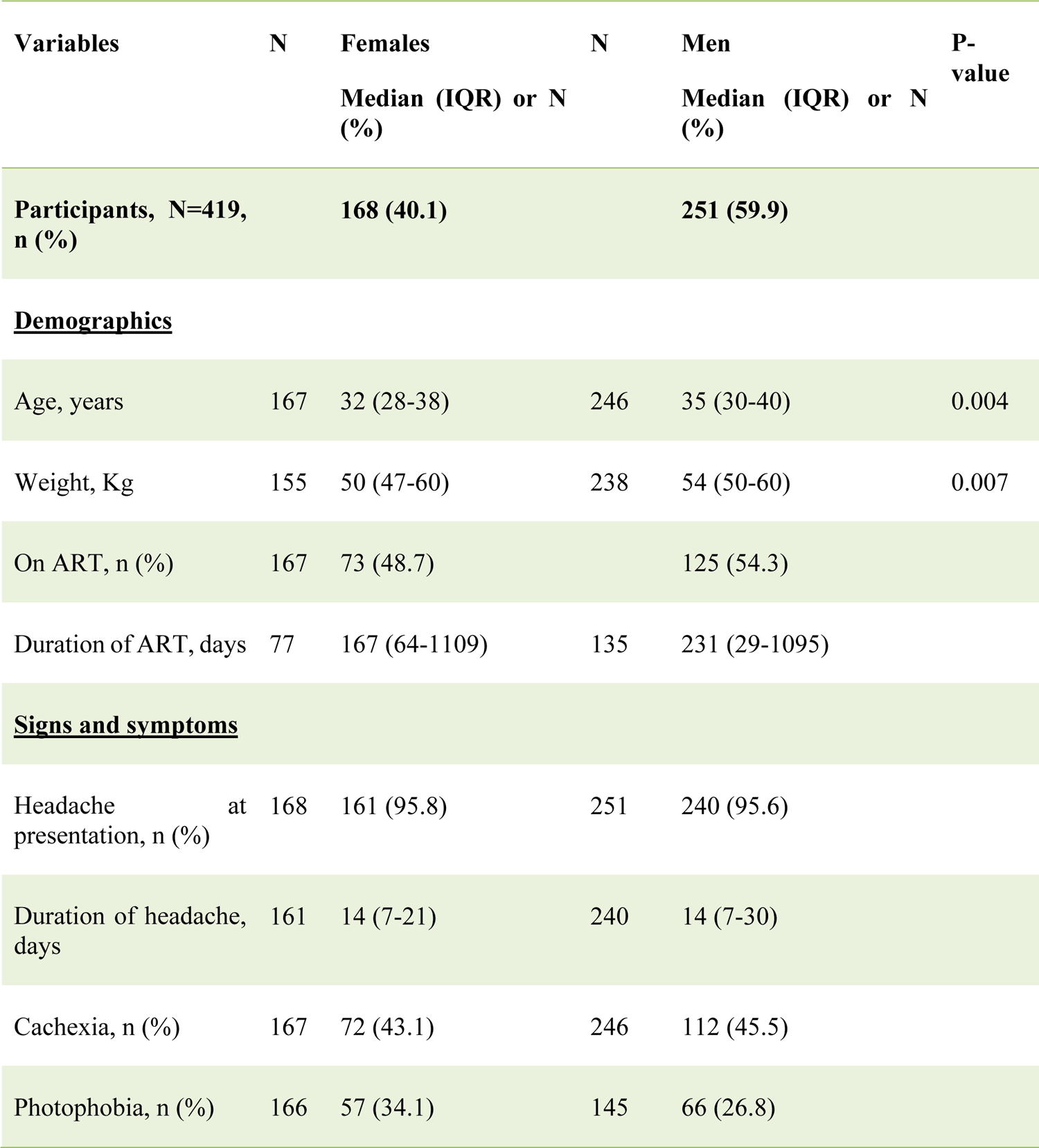
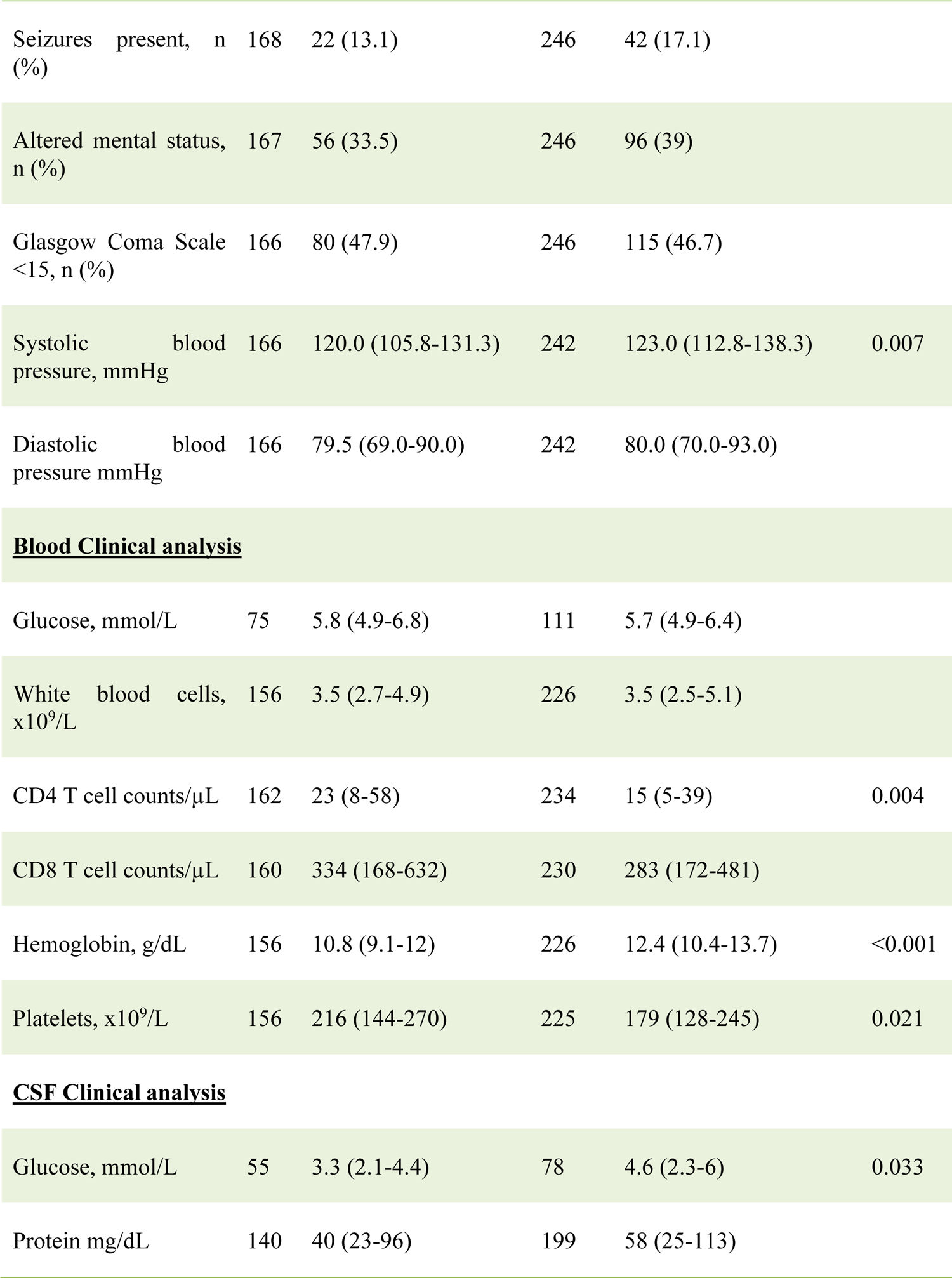
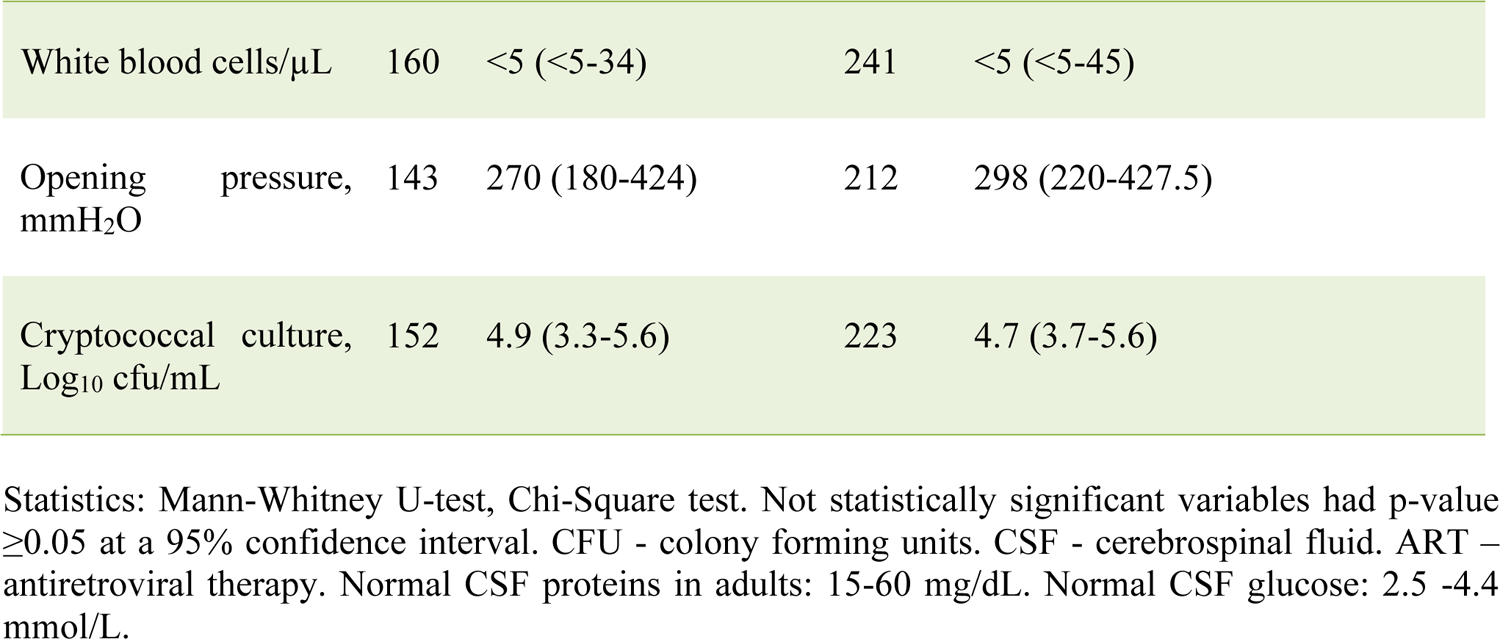
Baseline Demographics of People With HIV-Associated cryptococcal meningitis by Gender.

| Variables | N | Females<br>Median (IQR) or N (%) | N | Men<br>Median (IQR) or N (%) | P-value |
| --- | --- | --- | --- | --- | --- |
| Participants, N=419, n (%) |  | 168 (40.1) |  | 251 (59.9) |  |
| **<u>Demographics</u>** |  |  |  |  |  |
| Age, years | 167 | 32 (28-38) | 246 | 35 (30-40) | 0.004 |
| Weight, Kg | 155 | 50 (47-60) | 238 | 54 (50-60) | 0.007 |
| On ART, n (%) | 167 | 73 (48.7) |  | 125 (54.3) |  |
| Duration of ART, days | 77 | 167 (64-1109) | 135 | 231 (29-1095) |  |
| **<u>Signs and symptoms</u>** |  |  |  |  |  |
| Headache at presentation, n (%) | 168 | 161 (95.8) | 251 | 240 (95.6) |  |
| Duration of headache, days | 161 | 14 (7-21) | 240 | 14 (7-30) |  |
| Cachexia, n (%) | 167 | 72 (43.1) | 246 | 112 (45.5) |  |
| Photophobia, n (%) | 166 | 57 (34.1) | 145 | 66 (26.8) |  |
| Seizures present, n (%) | 168 | 22 (13.1) | 246 | 42 (17.1) |  |
| Altered mental status, n (%) | 167 | 56 (33.5) | 246 | 96 (39) |  |
| Glasgow Coma Scale <15, n (%) | 166 | 80 (47.9) | 246 | 115 (46.7) |  |
| Systolic blood pressure, mmHg | 166 | 120.0 (105.8-131.3) | 242 | 123.0 (112.8-138.3) | 0.007 |
| Diastolic blood pressure mmHg | 166 | 79.5 (69.0-90.0) | 242 | 80.0 (70.0-93.0) |  |
| **<u>Blood Clinical analysis</u>** |  |  |  |  |  |
| Glucose, mmol/L | 75 | 5.8 (4.9-6.8) | 111 | 5.7 (4.9-6.4) |  |
| White blood cells, x10<sup>9</sup>/L | 156 | 3.5 (2.7-4.9) | 226 | 3.5 (2.5-5.1) |  |
| CD4 T cell counts/μL | 162 | 23 (8-58) | 234 | 15 (5-39) | 0.004 |
| CD8 T cell counts/μL | 160 | 334 (168-632) | 230 | 283 (172-481) |  |
| Hemoglobin, g/dL | 156 | 10.8 (9.1-12) | 226 | 12.4 (10.4-13.7) | <0.001 |
| Platelets, x10<sup>9</sup>/L | 156 | 216 (144-270) | 225 | 179 (128-245) | 0.021 |
| **<u>CSF Clinical analysis</u>** |  |  |  |  |  |
| Glucose, mmol/L | 55 | 3.3 (2.1-4.4) | 78 | 4.6 (2.3-6) | 0.033 |
| Protein mg/dL | 140 | 40 (23-96) | 199 | 58 (25-113) |  |

|  |  |  |  |  |
| --- | --- | --- | --- | --- |
| White blood cells/μL | 160 | <5 (<5-34) | 241 | <5 (<5-45) |
| Opening pressure, mmH<sub>2</sub>O | 143 | 270 (180-424) | 212 | 298 (220-427.5) |
| Cryptococcal culture, Log<sub>10</sub> cfu/mL | 152 | 4.9 (3.3-5.6) | 223 | 4.7 (3.7-5.6) |

Statistics: Mann-Whitney U-test, Chi-Square test. Not statistically significant variables had p-value $\geq 0.05$ at a 95% confidence interval. CFU - colony forming units. CSF - cerebrospinal fluid. ART – antiretroviral therapy. Normal CSF proteins in adults: 15-60 mg/dL. Normal CSF glucose: 2.5 -4.4 mmol/L.

**Figure 1.**
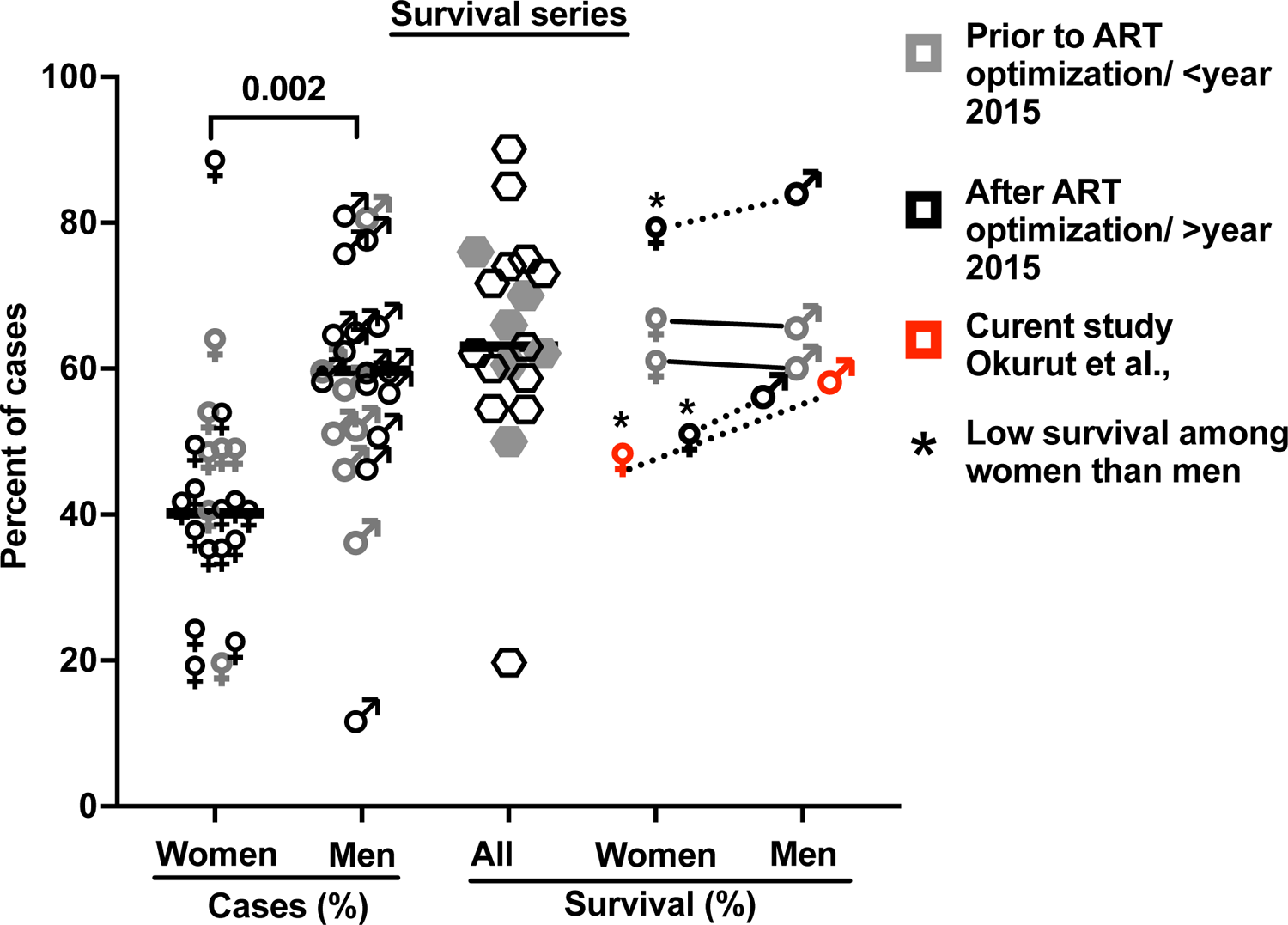
The proportion of cases with HIV-associated CM by gender and of related survival by gender in 21 published case series. These studies include 38,485 cases with 5,834 reported deaths, which accounted for a 15.2% case fatality rate (see Supplementary Table 1 for details and references ^7,9,19,26,28–44.^ Bars show median values. 5 studies report survival by gender (4 references and this report; Supplemental Table 1).

As anticipated, CD4^+^ T cell numbers in circulation were low, but were marginally higher in females than males, as were platelet counts and hemoglobin levels (**Table 1**). CSF protein was not consistently elevated, and WBC counts were low, despite a high burden of yeast. Each result was generally comparable by gender, except CSF glucose which tended to be lower among females than males (**Table 1**).

### 3.2 The 18 Weeks Survival on Antifungal Treatment

In the parent trial for this analysis, males and females from Uganda and South Africa were randomized 1:1 to receive sertraline (an antidepressant with putative antifungal activity) or standard treatment but showed no differences in survival between treatment groups at 18 weeks ^2^. An initial analysis by gender of an expanded data set, from which this current report is derived, showed significantly lower survival among 400 females vs. 577 males (at 50% vs. 57% survival, respectively) at 10 weeks, unadjusted (Hazard Ratio (HR) = 1.20; 95% CI, 1.00-1.45; p= 0.050) ^7^. These differences were greater yet in this current subset extended to 18 weeks of observation. Survival among females was 47% (71/150 females) vs. 59% among males (136/230 males) (Mantel-Cox proportional hazard ratio (HR)=1.4 (95% confidence interval, CI: 1.0–1.9); p=0.023) (**Figure 2**). Of note, survival was similar among males and females in two earlier studies reported prior to ART optimization (before 2015) but lower among females in two studies reported after ART optimization (after 2015), which include the current report (**Figure 1-2**). In the current study, survival overall, and by gender did not differ by ART experience (data not shown).

**Figure 2.**
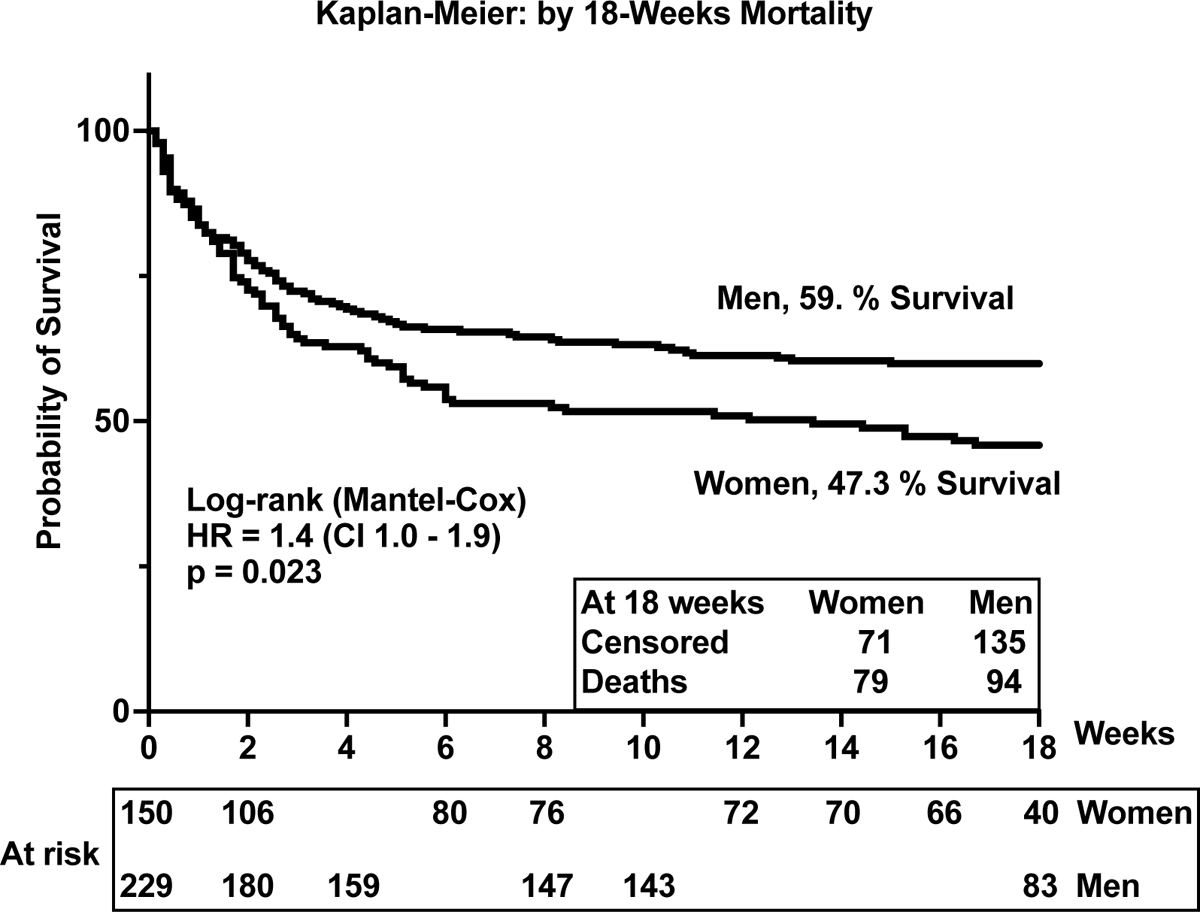
Survival among people with HIV-associated cryptococcal meningitis. The p-value <0.05 is statistically different.

Survival was similar by gender in the first two weeks of antifungal therapy (**Figure 2**). However, survival diverged thereafter, remaining consistently lower among females throughout the 18 weeks of observation. As noted, baseline demographics, signs and symptoms, blood and CSF analytes, and cryptococcal fungal burden were relatively similar among participants by gender and by survival (**Table 1**). Thus, we considered whether the concentrations and the composition of neuroimmune-induced factors at the site of severe cryptococcal disease in the CSF could underlie subsequent differences in survival by gender over time of observation.

### 3.3 Significant Differences in Neuroimmune Signatures in Cerebrospinal Fluid by Survival, Gender, and Intragender-Specific Survival

At baseline, we performed unsupervised Principal Component Analysis (PCA) as a primary approach to visualize the data variability and to explore potential unbiased differences in data clustering by gender, survival, and gender-specific survival (**Figure 3**). The PCA identified individual clusters that offered opportunities to structure downstream data analyses of the indicated model outcomes. The members in the cluster aggregated based on common attributes of the datasets showing high variability and distinct distribution of cytokines and chemokines between females and males (**Figure 3A**), between survivors and participants who died (**Figure 3B**), and within gender survival, among females (**Figure 3C**) and among males (**Figure 3D**). Eigenvector projections on principal components 1 and 2 (PC1 and PC2 respectively) indicated a high probability of neuroimmune variables predicting association with gender and survival outcome (**Figure 3 A-D**). High Eigenvalues >5 indicate the high capability of the selected covariables in predicting model-associated outcomes **(Table below Figure 3**). For each of the comparative groups, almost all showed such high Eigenvalues and, thereby, the separation between determinants in each group. These distinct patterns for PC1 and PC2 were closely correlated among participants by gender, subject by survival, and females by survival but less so for male survival in which 3 clusters were identified (correlation data not shown). Due to the striking data clustering observed in these groups, we next determined the specific neuroimmune factors contributing to the observed patterns using supervised univariate and multivariate data interrogation approaches.

**Figure 3.**
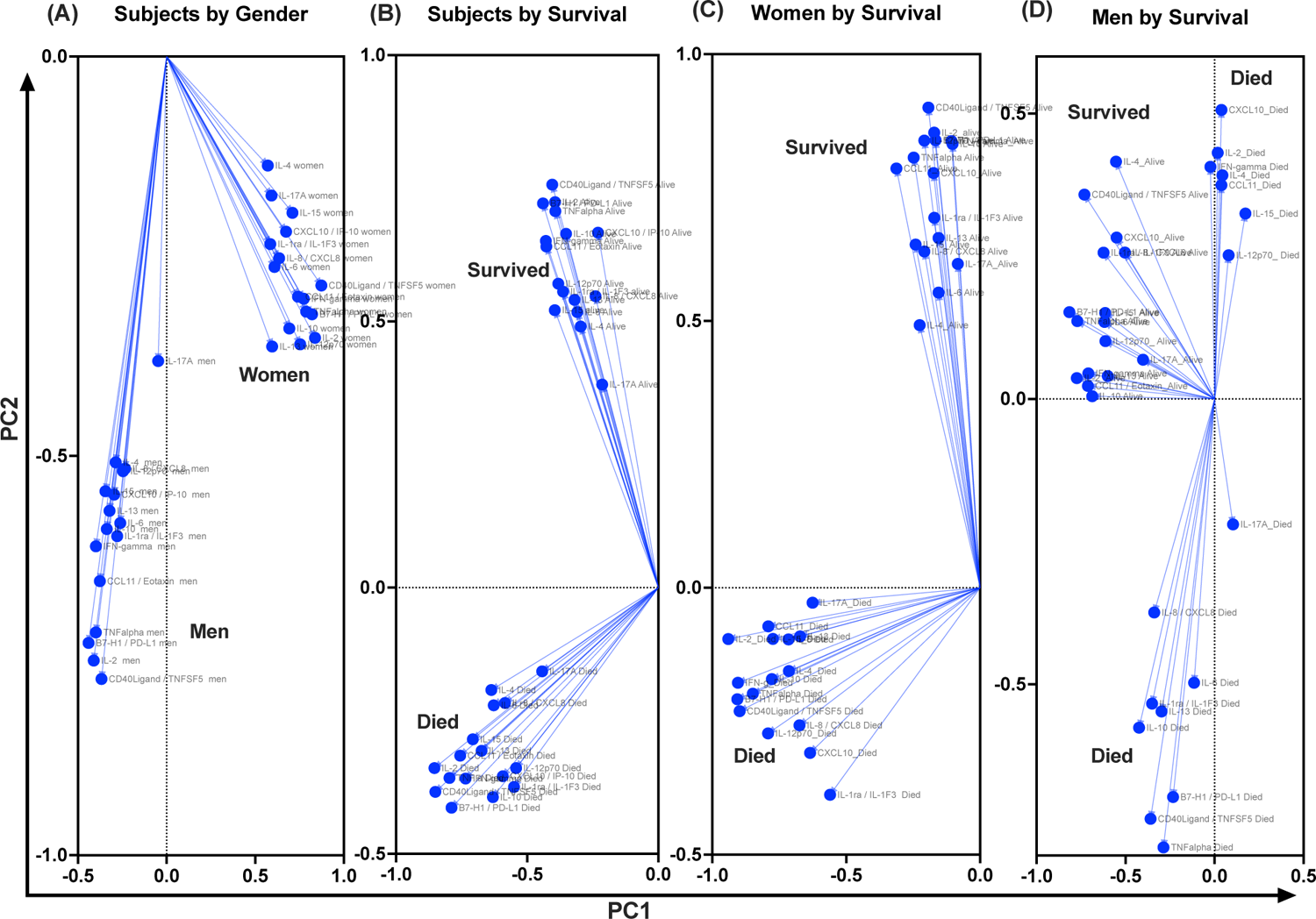
Projection of cytokine responses on Eigenvector correlation covariates on PC1 and PC 2 (axes) using unsupervised principal component analysis. Baseline cerebrospinal fluid (CSF) immune signature among participants who were diagnosed with HIV-associated CM showed distinct clusters by gender and by survival. PC - Principal component. Dots - show individual cytokines produced among participants projected on Eigenvector correlation covariates. Variables near the center (0; zero) are uncorrelated to the model and variables further away from the center (0; zero) are strongly correlated to the model. Variables among all cytokine projected on the variety of CSF cytokine patterns among participants by host survival (Figure 3A). The CSF secreted cytokines clustering by gender (Figure 3B). Cytokine clustering by female survival (Figure 3C). Cytokine clustering by men’s survival (Figure 3D).

**Adjacent Table to Figure 3:** PC1 shows the highest variance of loading on a single vector. PC2 shows the cumulative variance of loading that is orthogonal to PC1 with a center 0. Yet negative variables show the presence of hidden (latent) variables that can be only determined through inference using mathematical modeling or through direct measurement alongside observed variables (those with positive variance on PCA). The great the Eigenvalues than one (1) the greater the predictability power of the variable in determining the hidden or latent variance to the outcome.

CSF divergent-neuroimmune signatures by intragender-specific survival
| Group Analyzed | Participants by gender |  | Participants by Survival |  | Females by Survival |  | Males by Survival |  |
| --- | --- | --- | --- | --- | --- | --- | --- | --- |
| PC summary | PC1 | PC2 | PC1 | PC2 | PC1 | PC2 | PC1 | PC2 |
| Eigenvalue | 9.8 | 7.3 | 9.5 | 7.6 | 10.1 | 9.5 | 7.6 | 4.7 |
| Proportion of variance (%) | 30.6 | 22.7 | 29.7 | 23.9 | 31.7 | 29.7 | 23.9 | 14.6 |
| Cumulative proportion of variance (%) | 30.6 | 53.3 | 29.7 | 53.6 | 31.7 | 61.3 | 23.9 | 38.5 |

### 3.4 Innate Neuroinflammatory Cytokines in the Cerebrospinal Fluid Differ by Gender

The CSF of females with CM at baseline had significantly lower levels of selected innate cytokines than that of their male counterparts, particularly IL-1RA and IL-15, TNF-α and immune checkpoint, PD-L1 (all p<0.050) (**Table 2**). The remainder of the cytokines and chemokines interrogated did not differ significantly by gender but tended to be lower still among females than males (Table 2). We next considered whether such gender-specific differences were associated with the differences in survival by gender.

**Table 2.**
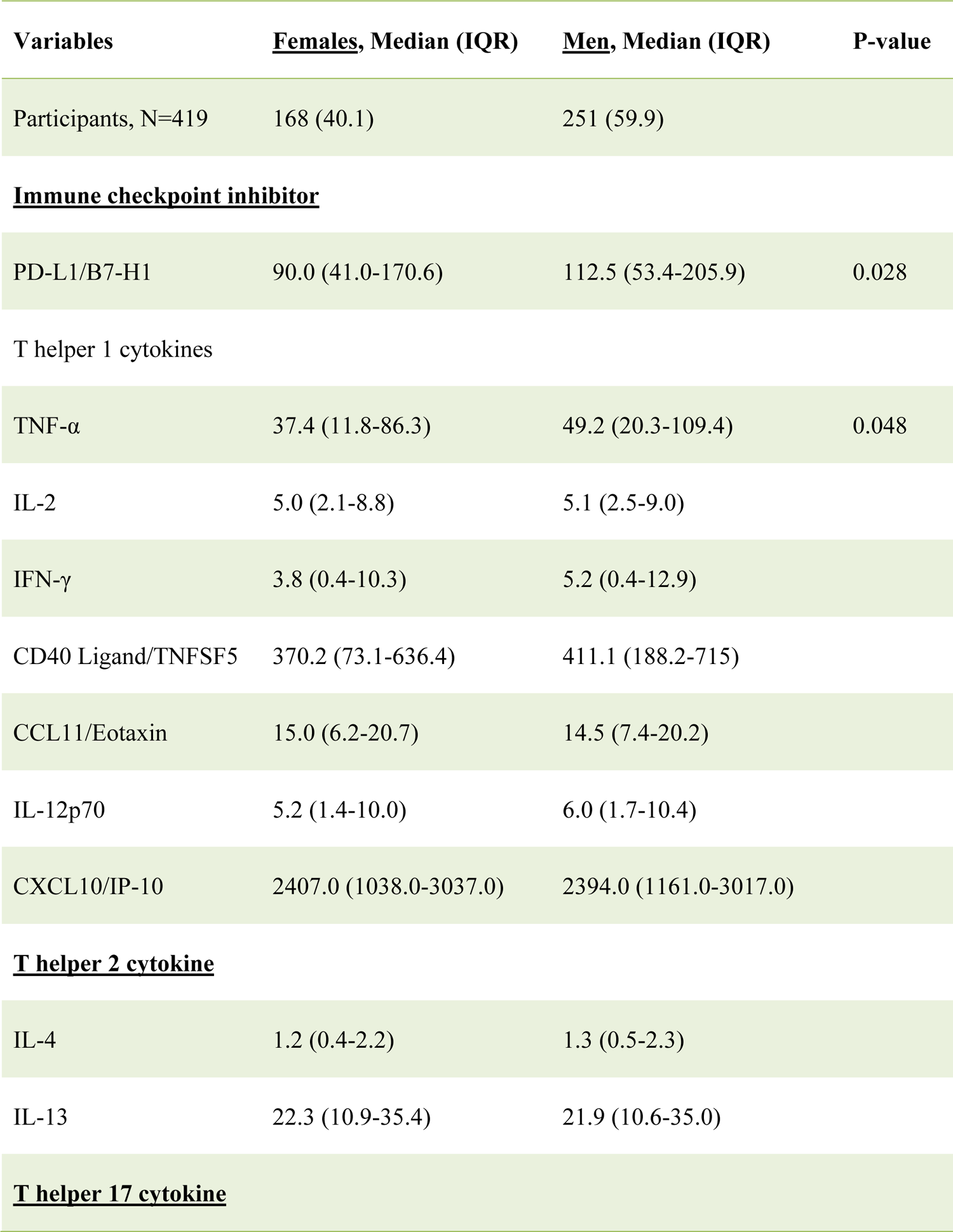
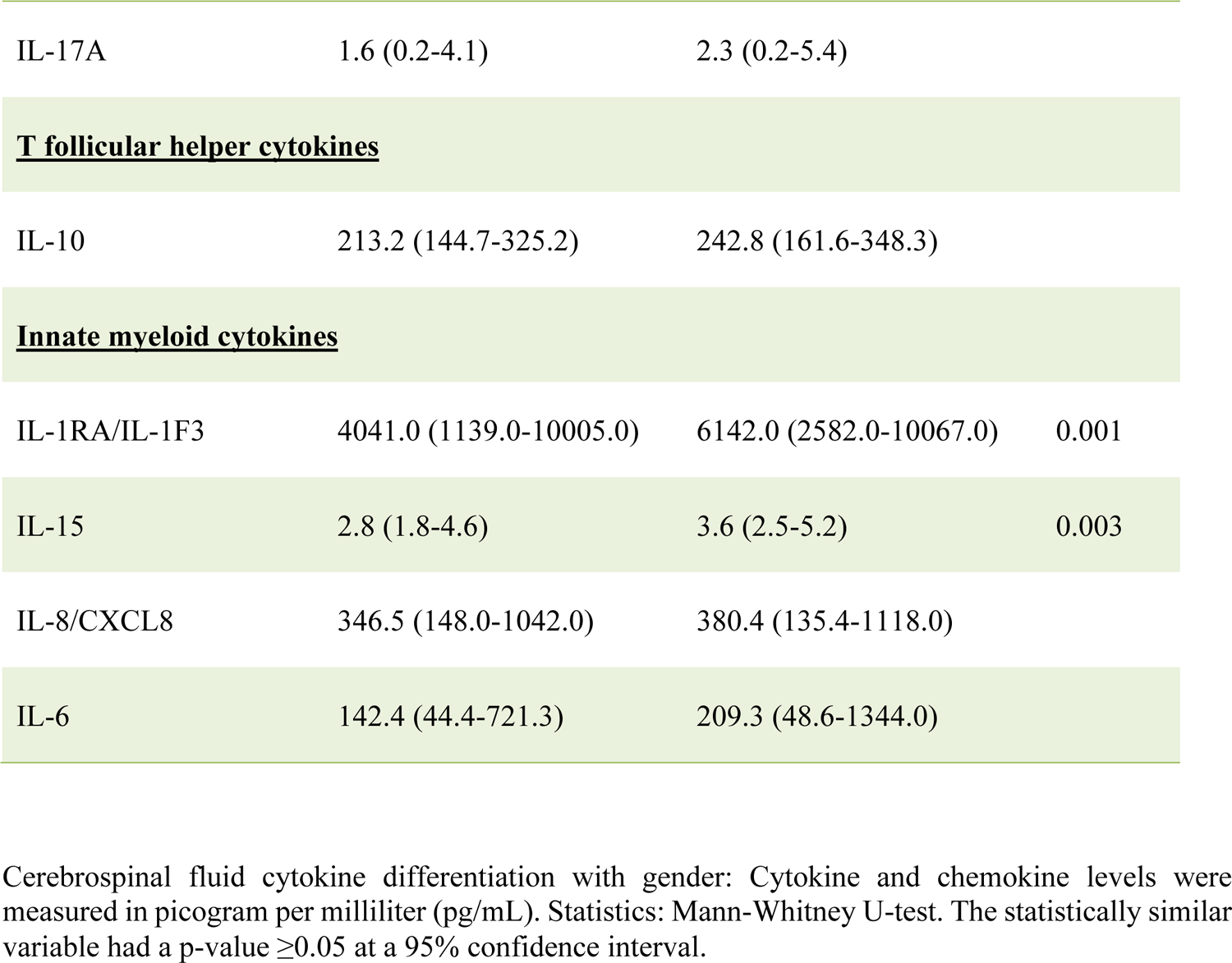
Differences in Baseline Immune Signature in Cerebrospinal Fluid Exudate by Gender Among Patients with HIV-Associated Cryptococcal Meningitis.

| Variables | <u>Females</u>, Median (IQR) | <u>Men</u>, Median (IQR) | P-value |
| --- | --- | --- | --- |
| Participants, N=419 | 168 (40.1) | 251 (59.9) |  |
| **<u>Immune checkpoint inhibitor</u>** |  |  |  |
| PD-L1/B7-H1 | 90.0 (41.0-170.6) | 112.5 (53.4-205.9) | 0.028 |
| T helper 1 cytokines |  |  |  |
| TNF-α | 37.4 (11.8-86.3) | 49.2 (20.3-109.4) | 0.048 |
| IL-2 | 5.0 (2.1-8.8) | 5.1 (2.5-9.0) |  |
| IFN-γ | 3.8 (0.4-10.3) | 5.2 (0.4-12.9) | |
| CD40 Ligand/TNFSF5 | 370.2 (73.1-636.4) | 411.1 (188.2-715) |  |
| CCL11/Eotaxin | 15.0 (6.2-20.7) | 14.5 (7.4-20.2) |  |
| IL-12p70 | 5.2 (1.4-10.0) | 6.0 (1.7-10.4) |  |
| CXCL10/IP-10 | 2407.0 (1038.0-3037.0) | 2394.0 (1161.0-3017.0) |  |
| **<u>T helper 2 cytokine</u>** |  |  |  |
| IL-4 | 1.2 (0.4-2.2) | 1.3 (0.5-2.3) |  |
| IL-13 | 22.3 (10.9-35.4) | 21.9 (10.6-35.0) |  |
| **<u>T helper 17 cytokine</u>** |  |  |  |
| IL-17A | 1.6 (0.2-4.1) | 2.3 (0.2-5.4) |  |
| **<u>T follicular helper cytokines</u>** |  |  |  |
| IL-10 | 213.2 (144.7-325.2) | 242.8 (161.6-348.3) |  |
| **<u>Innate myeloid cytokines</u>** |  |  |  |
| IL-1RA/IL-1F3 | 4041.0 (1139.0-10005.0) | 6142.0 (2582.0-10067.0) | 0.001 |
| IL-15 | 2.8 (1.8-4.6) | 3.6 (2.5-5.2) | 0.003 |
| IL-8/CXCL8 | 346.5 (148.0-1042.0) | 380.4 (135.4-1118.0) |  |
| IL-6 | 142.4 (44.4-721.3) | 209.3 (48.6-1344.0) |  |

Cerebrospinal fluid cytokine differentiation with gender: Cytokine and chemokine levels were measured in picogram per milliliter (pg/mL). Statistics: Mann-Whitney U-test. The statistically similar variable had a p-value $\geq 0.05$ at a 95% confidence interval.

### 3.5 Divergent Baseline Neuroimmune Cytokine Signature Predict Intragender Survival

#### 3.5.1 (i). Female Gender-Specific Survival Attributes

A number of relevant factors differed between females who survived or died during the 18 weeks of observation. The circulating CD4^+^ T cell numbers were generally very low, but median CD4^+^ T cells were 31 cells/µL vs. 14 cells/µL among females who survived vs. those who died (p=0.009) but did not differ by survival in males (data not shown). Using several alternative models, and after adjusting for cytokines (**Table 4 model 1)**, CXCL10 and CCL11 consistently predicted female survival but not in males. These soluble immune factors were consistently higher in magnitude among female survivors compared to those who died (**Table 3**). The levels of CXCL10 were significantly higher among females who survived than those who died, (p=0.013) (**Table 3**). By survival, CXCL10 levels in females who survived correlated with the number of CSF white cell counts (r=0.292, 95% CI: 0.067-0.488; p=0.010), but not in males **(Supplementary Figure 1)**. The fungal burden (CFUs) did not correlate with CXCL10 levels by either gender or gender-specific survival (data not shown).

**Table 3.**
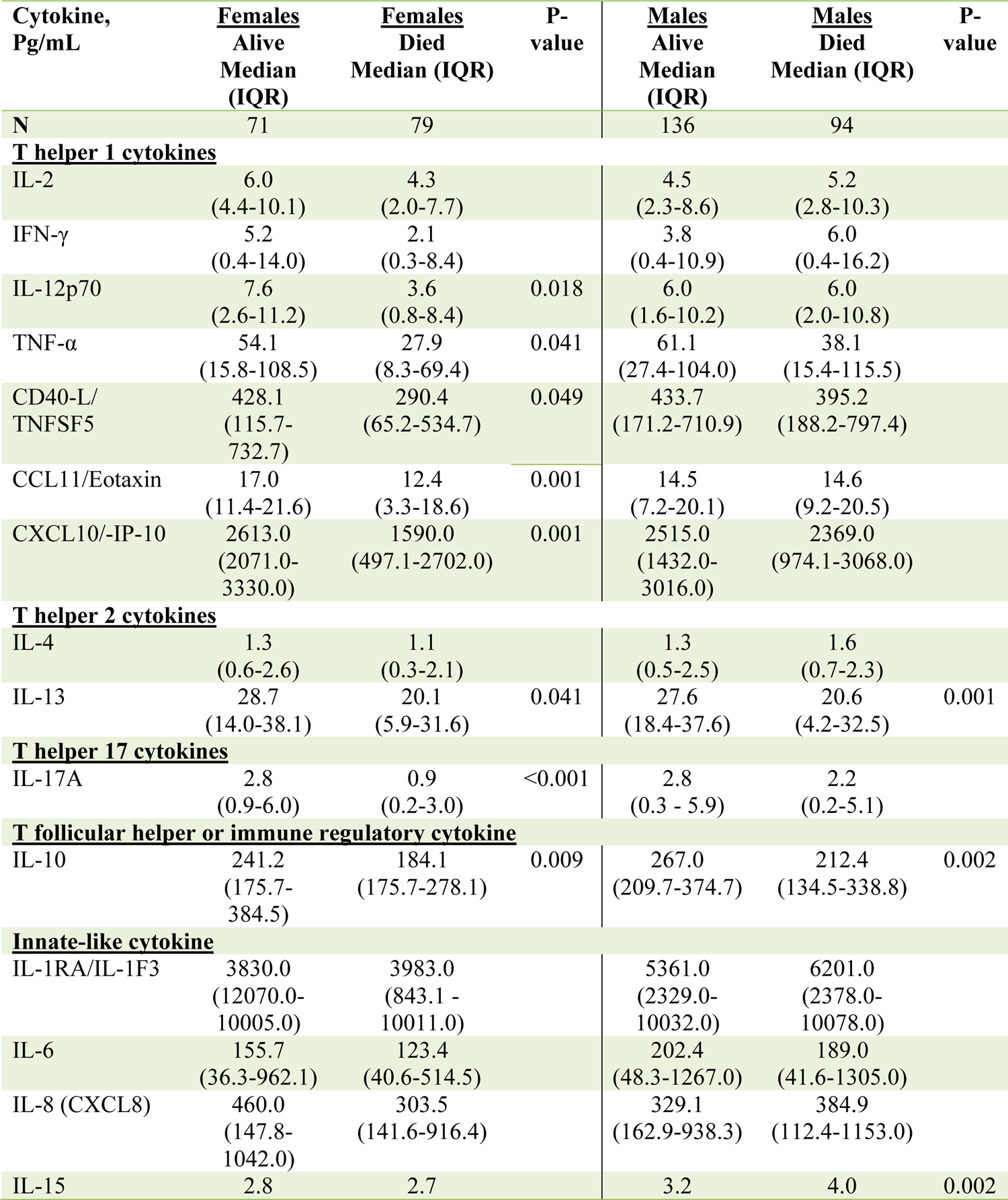
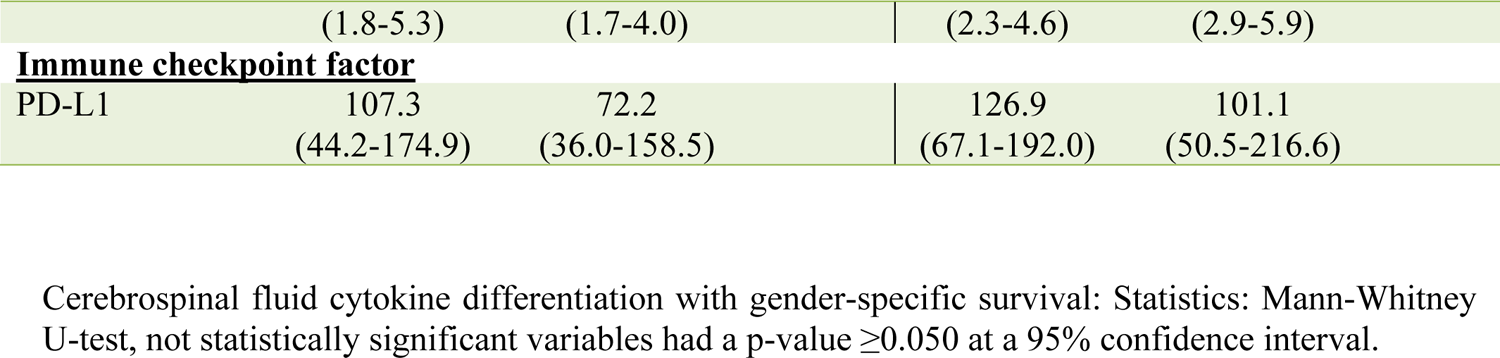
Univariate Difference in Baseline Levels of CSF Factors Associated with Gender Survival Among Patients with HIV-Associated CM.

**Table 4.**
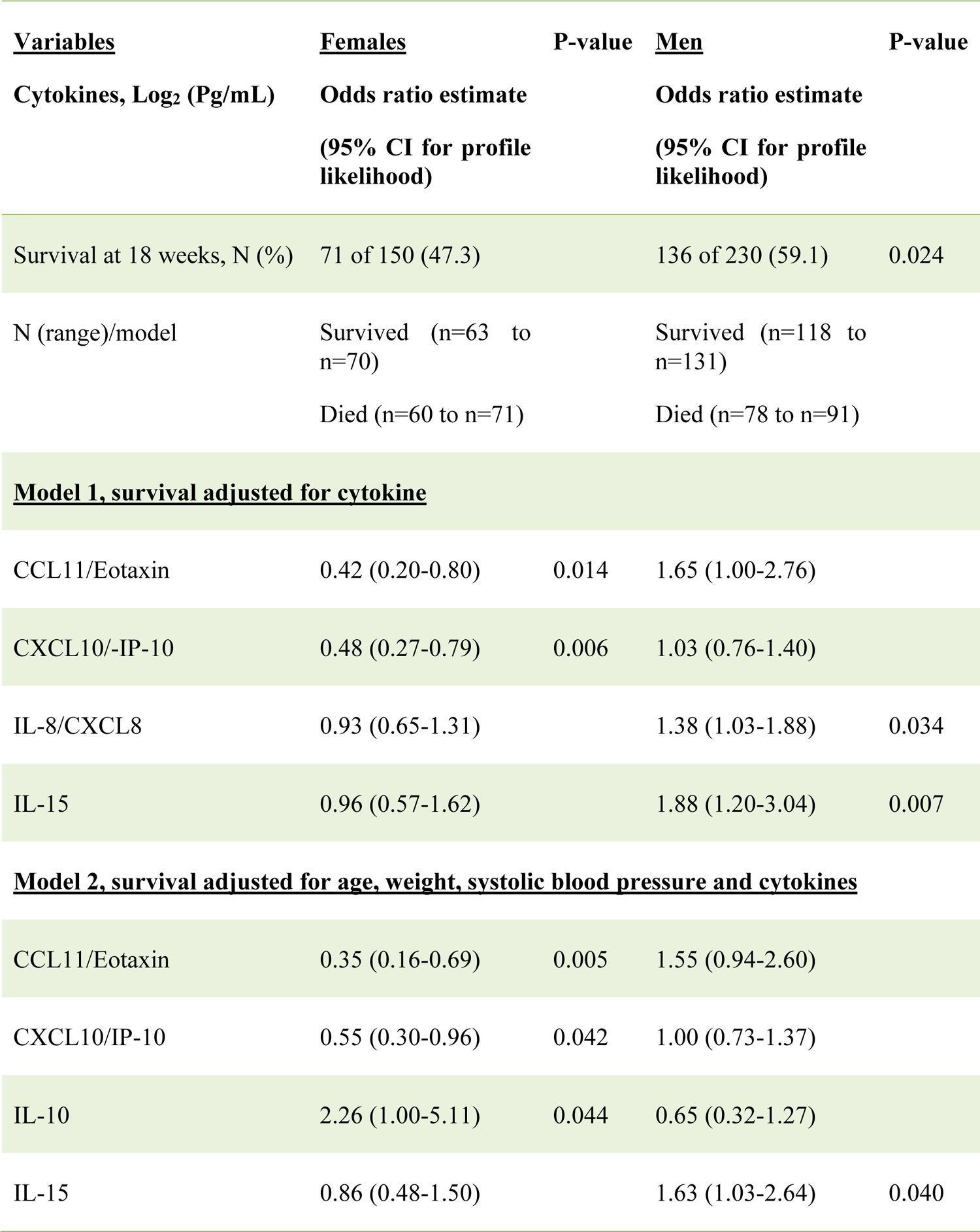
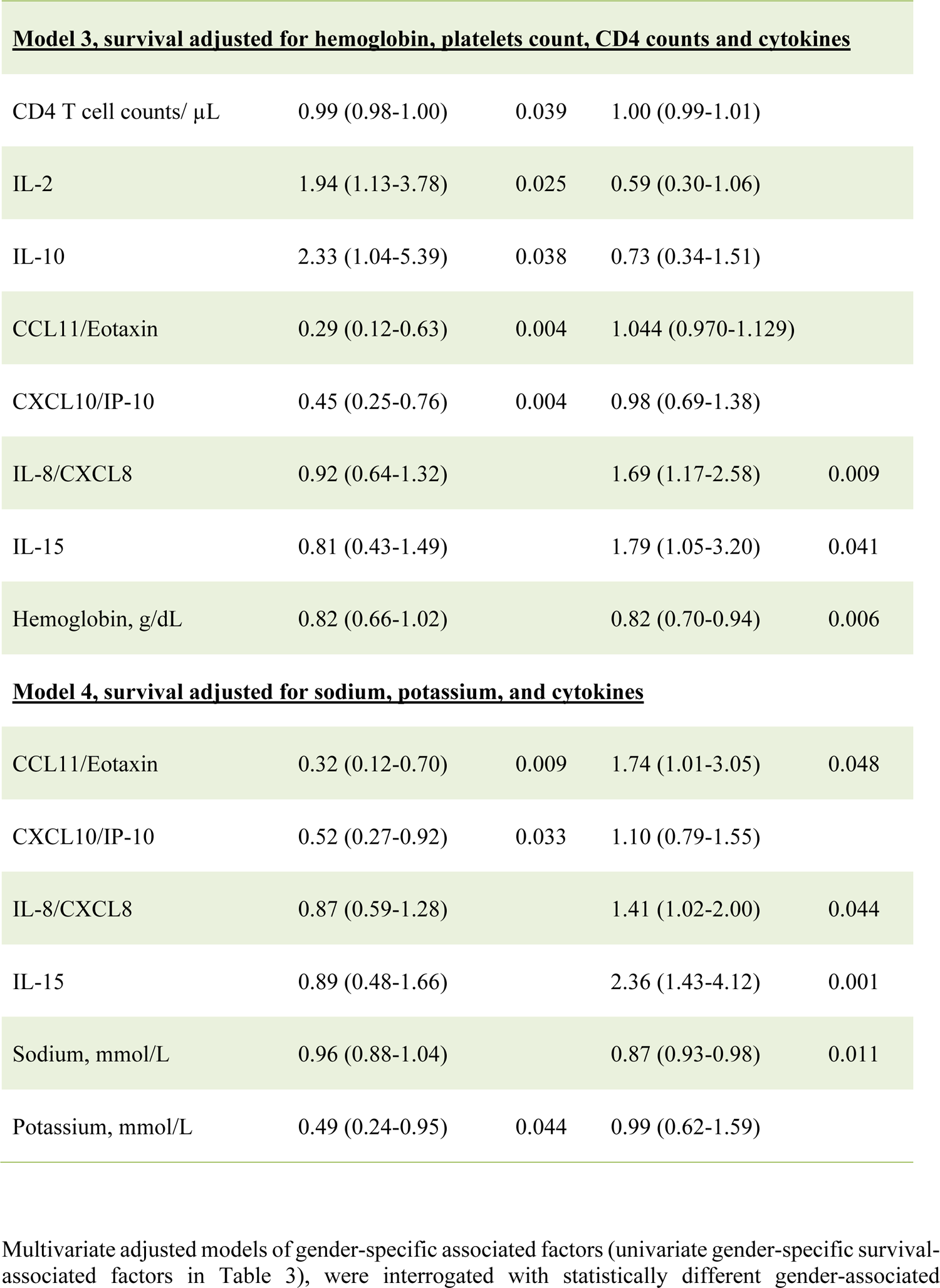
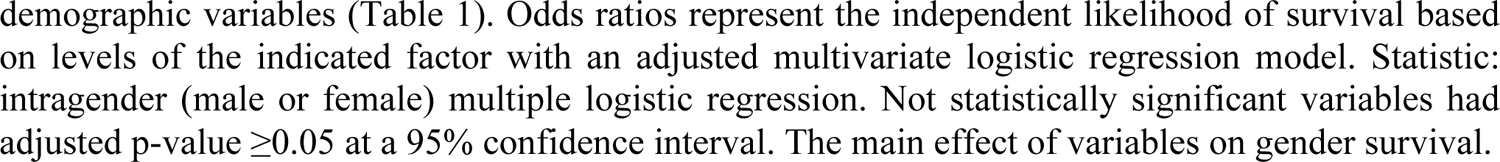
Baseline Independent Factors Predicting Intragender 18 Weeks Survival Among Patients with HIV-Associated Cryptococcal Meningitis on Antifungal Therapy.

The level of CCL11 (a chemoattractant produced by activated astrocytes, lymphocytes, and macrophages), was significantly higher in female survivors than in those who died, but not among males (**Table 3**). After adjusting for other cytokines, higher CCL11 expression still predicted female but not male survival (**Table 4, Model 1).** T cell-related factors (e.g., IL-12p70, IL-17A, and IFN-γ) in the univariate models were also consistently increased in females who survived compared to those who died, but not in males (**Table 3**). The levels of other cytokines and the neutrophils chemokine IL-8 showed no difference among females by survival (**Table 3**). Only the IL-10 level was increased in both genders among those who survived in univariable analysis (**Table 3**). However, after adjusting for other factors, the levels of IL-10 expression were correlated with only female survival, but not with male survival (**Table 4, Models 2 and 3)**, as was IL-2 (**Table 4, Model 3)**.

In summary, although immune parameters were lower among all females vs. males, the females who survived showed consistently elevated levels of both myeloid-derived chemokines (CCL11), lymphoid-derived chemokine (CXCL10), T cell-derived cytokines, and the regulatory molecule PD-L1, the differences that were mostly distinct to survival in females, but not in males **(Supplementary Figure 1)**.

#### 3.5.2 (ii). Male Gender-Specific Survival Attributes

Males who survived had significantly higher hemoglobin compared with those who died, even after adjusting for the platelet counts and CD4+ T cell counts (**Table 4, Model 3).** Males, but not females, who survived had significantly lower IL-15 than those who died (**Table 3),** even after adjusting for other cytokines (**Table 4, all Models).** The IL-15 levels were independent of fungal burden (CFUs) and CSF white cell counts (data not shown). After adjusting for other factors, lower levels of the neutrophil chemoattractant IL-8 also consistently predicted male survival but not among females (**Table 4, Models 1, 3-4).** Unlike in males, neither low levels of IL-15 nor IL-8 were associated with survival in females. As noted above, elevated levels of regulatory IL-10 were associated with survival in both genders but did not correlate with male survival **(Supplementary Figure 1)**.

## 4 DISCUSSION

In a large cohort of adults with CM in Uganda on anti-fungal therapy, survival among females was significantly lower compared with that of their male counterparts. Evaluating gender-specific mortality is uncommon in this context (only 5 including the current 21 studies; Supplemental Table S1), Unlike two previous studies which showed no difference in mortality by gender reported ^26,44^, the two studies showing such a differences ^7,42^ were performed in people treated with antifungal therapy, which helps control fungal burden. Combining antifungal drugs with immunomodulatory interventions could limit the fungal assault on the CNS, illustrated by brain lesions from magnetic resonance imaging and from autopsy examinations among cases with neurological focal infections ^62–65^. With appreciable control of fungal burden, immune responses may be the more prominent determinant of clinical outcomes.

We have identified distinct immune-biological differences in CSF by gender and gender-specific survival. The signatures associated with survival in females are distinctive from those in males. These differences were independent of baseline clinical features and cryptococcal fungal burden (CFUs). Most consistent in both the univariate and in multivariate gender-associated survival predictive models were the lower levels of CSF CXCL10/IP-10 and CCL11/Eotaxin that distinguished females who died from survivors, but not males. In contrast, high levels of IL-15 and IL-8 differentiated males who died matched to those who survived, but not females. Several T cell cytokines (IFN-γ, TNF-α, IL-13, and IL-17A) similarly exhibited diminished levels of expression in females who died matched survivors, but not in males. Lower levels of immune regulatory IL-10 expression were linked with an increased threat of death in both genders. Thus, the biological significance of gender-specific CSF immune signatures suggested among participants with CM may underlie important immune-based mechanisms indispensable to improving antifungal treatment and survival predominantly in women with demonstrated distinct survival-associated cytokine and chemokine patterns.

Consistent with the univariate model observations, improved survival at 2-10 weeks was associated with elevation in the levels of soluble CSF cytokines Th1 IL-2, IFN-γ, TNF-α, Th2 IL-4, and Th17 IL-17A, together with innate IL-6 cytokine ^66,67^. Soluble T-lymphokines levels in CSF are proposed to derive from CXCL10 ^68^ and CCL11 ^69^ mediated stimulation of CNS resident immune cells. Moreover, the use of cryptococcal antigens (such as GXM) to stimulate peripheral immunocytes demonstrated, that patients with improved 10 weeks survival had increased expression of these cytokines among polyfunctional differentiated CD4 T cells ^67^. Other chemokines demonstrated to predict improved survival included the elevated levels of monocytes chemotactic protein-1 and macrophages inflammatory protein-1 ^66^. Despite demonstrated correlates of survival in the peripheral circulation ^70,71^, the importance of characterizing immune responses in the CNS, at the foci of infection, cannot be underestimated since CNS responses in larger studies do not often correlate with those in the peripheral circulation ^70,71^. The disparity between peripheral and CNS observed responses is a characteristic indicator of local mechanisms that influences responses. Compartment-specific responses have been noted previously ^19,20^ and argue that we should be investigating the response in CSF. Thus, the current study is unique in characterizing the distinctive neuroimmune signatures defining survival by gender in CNS, at the foci of disease.

In theory, the microbial invasion of the CNS activates resident immune cells to produce neuroimmune cellular activating cytokines, chemoattractant chemokines, and surrounding tissue basal cellular immune mediators (prostaglandins, leukotrienes, et cetera). Diverse effector cells include resident microglial, astrocytes, oligodendrocytes, CNS surveillance phagocytes (monocytes, macrophages, neutrophils), dendritic cells, adaptive T and B cells, innate (natural killer cells), and basal barrier epithelial cells ^72^. Other cells may be drawn by extravasation and diapedesis through the protective vascular barriers to infiltrate the CNS in response to infection and/or to ensuing meningoencephalitis ^19,20^. The consistent use of antifungal therapy in this study helps to control fungal replication and the burden of fungi ^2,9,26,73^. With relatively effective antifungal therapy modeling immune factors highlights the role of the immune system in determining immune-mediated damage vs. protection in the CSF, and ultimately disease outcome ^67,74,75^.

Although cryptococcal fungal burden and other clinical parameters in blood or in CSF did not differ by gender or gender-associated survival, increased production of IL-1RA, IL-15, and PD-L1 in the CSF of males overall may have influenced their poor survival than in their female counterparts. Lower male but not female survival was associated with high levels of myeloid cytokines, including the pleiotropic IL-15 and neutrophil-activating factors IL-8, which may influence poor survival in males by the distinctive mechanisms. In literature, IL-8 expression derived from microglia in CNS in response to neuroinflammation ^76,77^, from monocytes and autocrine IL-8 production by neutrophils enhances immune activation and cellular proliferation ^18^. The elevated intrathecal IL-15 and IL-8 in the CSF can enhance neutrophils-induced NETosis (Neutrophils traps). The increased neutrophil trap formation may propagate thrombotic vasculitis, cryptococcomas, and fungal occlusions in subarachnoid ventricular and arterial spaces in those with poor survival. Together IL-15 and IL-8 elevation can increase local inflammation in the brain, leading to vasculitis in subarachnoid spaces, interfering with blood supply, promoting ischemia, necrosis, and infarction of the brain tissue, and potentially leading to increased risk of death. Thus, the relatively high levels of IL-15 expression among males who died and the inverse relationship of IL-15 levels with those of immune regulatory IL-10 and immune checkpoint PD-L1 among males who died highlights the dynamic functional interrelationship of the potential loss of IL-15 immune regulatory function in influencing the greater risk of death among patients presenting with this neuroimmunological phenotype. Low levels of IL-15 may limit the mitotic division of especially fungal-infected macrophages which may attenuate the propagation of endogenous pathogens in quiescent cells ^18^. Thus, the observed low levels of IL-15 and IL-8 among male survivors may limit intracellular cryptococcal fungal replication potentially leading to low fungal burden and improved survival.

Among neutrophils activated factors that shape neuroimmune response and outcome in CNS infection and in sequelae include observations that neutrophil induced responses exacerbate tissue injury through self-directed host and pathogen-mediated mechanisms ^78,79^. That neutrophil activation is associated with the propagation of tissue necrosis, hypoxia, and nutritional supply impairment especially among people with sepsis and in those with multiple organ failure, that underlie brain injury ^78,79^. In this context, activation of neutrophil recruiting cytokines and chemokine, at the foci of infection, in the CNS is detrimental to the host ^76,78^, leading to the release of toxic granules, that are destructive to connective tissues, leading to vasculitis with tissue necrosis, infarction, hypoxia, and potentially death. Indeed, in cryptococcal meningitis, neutrophilia in circulation was associated with poor survival outcome ^80^, brain hypoxia ^15,81,82^, subarachnoid blood vessel occlusion ^83^, brain tissue necrosis, and cerebellar infarction ^64^. Subsequently, survivors of cryptococcal meningitis suffered long-term CNS abnormalities associated with impaired faculty observed in sequelae among people without HIV infection ^62,84^. The persistent CNS abnormalities in sequelae noted in humans with fungal infections were similar to those observed among mouse models of Alzheimer’s disease ^85^. In other fatal meningitis focal infections including tuberculosis meningitis and bacterial meningitis, neutrophils account for the majority of cells in the CSF ^86,87^. However, neither we nor others have characterized in detail the neutrophil myeloid lineages and role among CSF white cells in the setting of CM-associated survival.

Among key differences in female intragender survival with cryptococcal meningitis, survivors tended to shade into CSF relatively high amount of neuroimmune mediating soluble cytokine and chemokine responses across the panel. The higher levels of Th1 IL-2, IFN-γ, TNF-α, IL-12p70, Th17 IL-17A, and immune regulatory IL-10 may influence survival in females by distinct CXCL10 and CCL11 chemokine-stimulated mechanisms. In contrast to females who died, females with higher levels of the myeloid chemokine CCL11 and the lymphoid chemoattractant CXCL10 were more likely to survive, but not men. CXCL10 produced by astrocytes and microglia cells or by CNS resident monocytes, macrophages, and dendritic cells stimulates Th1 cytotoxic CD8 T cells, immune modulatory CD4 T cells, regulatory T cells, NK and regulatory B cells, and regulatory T cells via its CXCR3 receptor-mediated stimulation ^88^. The cytotoxic response to infected macrophages is potentially enhanced in the presence of elevated chemokine in the central nervous, because infected cells respond to chemoattractant proteins at the site of infection ^89,90^, whereas immune regulatory cells can modulate inflammation and cellular activation to stimulate tissue recovery ^91^. Thus, independent of the baseline CSF cryptococcal CFUs and the baseline clinical features, the CCL11 and CXCL10 balance is critical in immune regulation of the Th1/Th2/17 balance, modulation of CNS cellular influx that is important in better survival of host with HIV-associated cryptococcal meningitis.

Interleukin (IL)-12 is observed as an important factor in the immune control of *Cryptococcus* meningitis as it is observed to influence mechanisms that inhibited cryptococcal fungal replication among *Cryptococcus*-infected macrophages in vitro ^92^. In mouse models, early preemptive use of IL-12 as an adjuvant promoted survival of mice in early use after cryptococcal fungal ^92^. Whereas the late treatment of mice with IL-12 after the establishment of cryptococcal infection resulted in more mice dying from fungal infection-attributable deaths ^92^. In the current study, IL-12p70 was elevated among female survivors but not males. In the CNS IL-12 is produced by dendritic cells, and is observed to regulate T cell, and NK cell function, and influence the differentiation of the Th1 cellular phenotype^93^. With the prominence of T cells and B cells, in the CSF with cryptococcal meningitis ^19,20^, increased IL-12p70 shading into the CSF can enhance fungal-specific immunocytes activation to mediate fungal neuroimmune mechanisms for control and for improved host survival.

The Th17 cytokine, IL17A, modulated through RORγt regulatory pathway was observed to work in tandem with Th1-modulated cytokines to regulate neuroimmune responses induced to CNS infections ^94^. In particular, IL-17A activates Th17 immunocytes in response to tissue-resident infection. In cryptococcal model infection, IL-17 promoted the development of majority giants cryptococcal cells that have limited ability to cross the blood-brain barrier, leading to localized cryptococcal fungal infection with limited spread to the brain parenchyma where it would cause lethal disease ^65,95^. Among females, IL-17A was upregulated among those with better survival outcomes. Other Th1 cytokines upregulated among females survivors included IFN-γ, TNF-α and their immune activating factors, CD40 ligand that was demonstrated to facilitate the formation of reactive phagosomes, activation of reactive oxygen and nitrogen species that mediate endogenous killing mechanisms of *Cryptococci* ingesting macrophages ^65,95^. Collectively, these immune activating complementary factors, were each increased with increased survival among females. And in the aggregate, these novel findings (reported for the very first in the setting of CM gender-specific survival), drive the question as to how these results may be harnessed for targeted gender-specific therapy to optimize survival where fatal outcomes seem to be propagated by immune-mediated mechanisms that appear to operate independent of the fungal burden, clinical presentation, and current antifungal therapy.

## 5 CONCLUSION

Survival from HIV-associated CM remains significantly lower in females. We found novel distinctive and divergent intragender-associated CSF cytokine signatures at the time of clinical presentation that was independent of the baseline fungal burden and clinical features. Among females, increased concentration of CCL11, CXCL10, and T cell subset associated cytokines mediating the Th1 and th17 fungal immunopathogenesis pathways to be divergently augmented with female survival but not in males. However, among males, low levels of the neutrophil-activating chemokines IL-8 and the pleiotropic cytokine, IL-15 potentially fashion antifungal regulation neuroimmune milieu, distinctively expressed with improved male survival but not in females. Increased expression of IL-10 and PD-L1 regulatory mechanisms accompany improved survival in both genders identifying common features of a successful CSF profile. The elevated induction of CCL11, CXCL10, IL-17A, and IL-12p70 neuroimmune cytokine signature by female intragender-specific survival in the CSF and their strong positive correlation to IL-10 and PD-L1 highlight the interdependence of these responses in shaping immune homeostasis and survival associated-neuroimmune modulating milieu. These key observations reveal promising signals to consider the impact of more robust responses in females, particularly T cell-dependent (CXCL10, Th1/Th17 cytokines), and lower myeloid responses in males (e.g., IL-15 and IL-8) and their regulators in controlling outcomes. This data point to the potential neuroimmune mechanistic targets which could be modified to improve treatment and clinical outcomes in those with reversible deaths with immune-based combination antifungal therapy.

## 6 LIMITATIONS

One limitation is that the cross-sectional, exploratory study design generated data could not explain marker and clinical variations after the baseline and prior to death outcome or censor, which might be key to explaining the factors that underlie the observed gender-survival subjective mechanisms. The differential of myeloid vs. lymphoid white cells in the CSF compartment was largely unavailable. The limitations of our interpretations of the baseline cross-sectional findings could be enhanced with longitudinal data to appropriately capture and model changes in the immune profile from diagnosis to eventual survival or death.

## 8 AUTHOR CONTRIBUTION

SO, EO, DBM, ENJ, YCM, DRB, JR, OOJ, HK, BB, and FB, conceptualized the study, framed the hypothesis, and edited the manuscript. SO, YCM, DRB, JR, DBM, and ENJ sourced for funding, designed experiments, guided data analysis, and drafted the manuscript. SO, performed statistical data analysis, compiled the results, put together the first drafted manuscript, and managed the review and publication process. The authors read and approved the final version of the manuscript for online publication.

## Data Availability

Data is available on request from authors.

## ACKNOWLEDGEMENTS

We appreciate the study participants for their involvement in the parent study. We thank the ASTRO trial team for the clinical management of patients and for data collection including Abdu Musubire, Lilian Tugume, Jane Francis Ndyetukira, Cynthia Ahimbisibwe, Florence Kugonza, Alisat Sadiq, Radha Rajasingham, Catherine Nanteza, Kiiza Tadeo Kandole, and Darlisha Williams, Edward Mpoza, Fiona Cresswell, Andrew Akampurira, Apio Alison, Caleb Skipper, Kenneth Ssebambulidde, and Mahsa Abassi. We thank the institutional contribution from Infectious Diseases Institute from Bosco Kafufu, Andrew Kambugu. We thank the PhD program mentorship support received from; Infectious Diseases Institute Capacity Building Unit especially from Barbara D. Castelnuovo, Aidah Nanvuma, Stephen Okoboi, and the Statistics unit, especially from Agnes Kiragga. John Hopkins University, School of Medicine and Bloomberg School of Public Health, Department of Molecular Microbiology, and Immunology, especially from Robert R. Bollinger and Arturo Casadevall. University of Colorado, Denver, Anschutz Medical Campus Aurora, Division of Infectious Disease, Department of Medicine and Veteran Affairs Research Services especially from Brent Palmer, Tina Powell, and Jeremy Rakhola. We thank the statistical data analysis mentorship received from the University of Minnesota, especially from Ananta S. Bangdiwala and Kathy Hullsiek Huppler.

## 10 ETHICAL CONSIDERATIONS

Participants provided prospective informed written and signed consent and storage consent for specimen use in future studies. Makerere University Higher Degrees Research and Ethics Committee of the School of Biomedical Sciences granted a waiver of informed consent and ethical approvals to analyze the data. The protocol was registered and approved by the National Council of Science and Technology. The parent clinical trial was approved by the National Drug Authority and by the Uganda National Council for Science and Technology. The patient’s identifiers were alphanumeric coded and could not identify those patients during the data search.

## 11 CONTRIBUTIONS TO THE FIELD

The study involved a large sample size with equal ratios between comparative variables that give strong confidence to weighted treatment outcome predictors of HIV-associated cryptococcal meningitis. We discovered the critical role of biological gender in the assessment of survival among patients with HIV-associated cryptococcal meningitis with discrepant bias in women’s poor survival. And that immune response differs within gender-specific survival. Together we highlighted the potential role of neuroimmune mechanisms in contributing to the differences in biological gender in survival. The observed immune mechanism illustrates the potential ability of the underlying stimulus in advancing immune-based treatment to work in synergy to improve the efficacy of antifungal drugs to improve survival among preventable deaths.

Interim, we propose close immune monitoring of CCL11/Eotaxin, CXCL10/IP-10, IL-12p70, and IL-17A as key indicators of the beneficial Th1 neuroinflammatory cytokine milieu among HIV and cryptococcal meningitis diagnosed women. We recommended mechanisms to augment CCL11/Eotaxin, CXCL10/IP-10, IL-12p70, and IL-17A cytokine deficient among women in addition to interventions to holistically refurbish the dysregulated or exhausted immune response among those co-infected in an attempt to significantly improve women’s survival outcome. We propose close monitoring of IL-15, IL-8/CXCL8 together with IL-10 and PD-L1/B7-H1 dysregulation as key indicators of men’s poor survival leading pathway. We recommend mechanisms to restore IL-15 and IL-8/CXCL8, IL-10, and PD-L1/B7-H1 checkpoint responses to improve men’s survival outcomes. It is plausible that switching the CCL11/Eotaxin and CXCL10/IP-10 chemokine balance in later stages of disease progression to a potentially myeloid pathway can lead to a significant self-reliant neuro-injury inflicting pathway leading to poor survival outcomes.

## 1. SUPPLEMENTARY MATERIALS

**Supplementary Figure 1.**
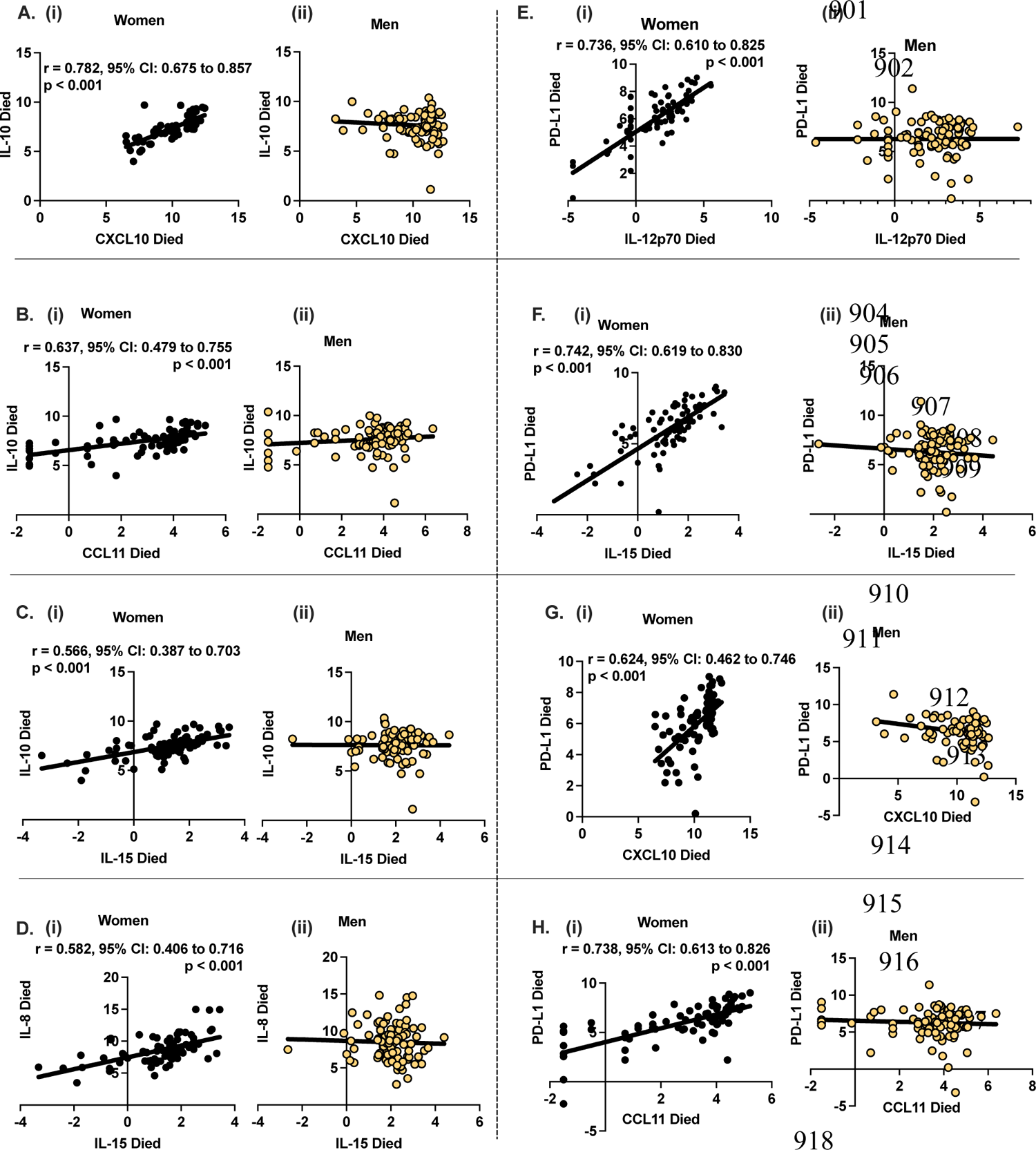
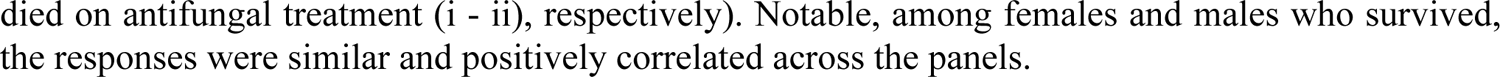
Spearman’s correlation of regulatory elements IL-10 and PD-L1 by gender among multivariate predictors of gender-specific survival among females and males who died on antifungal treatment. A-C correlation of IL-10 with A - CCL11/Eotaxin, B - CXCL10/IP-10, and C - IL-15 among females and males who died on antifungal treatment, (i - ii, respectively). D – correlation of IL-8 with IL-15 among females and males who died on antifungal treatment. E-H – correlation of PD-L1 with E – IL12p70, F – IL-15, G – CXCL10, and H – CCL11 among females and males who died on antifungal treatment (i - ii), respectively). Notable, among females and males who survived, the responses were similar and positively correlated across the panels.

**Supplementary Table S1.**
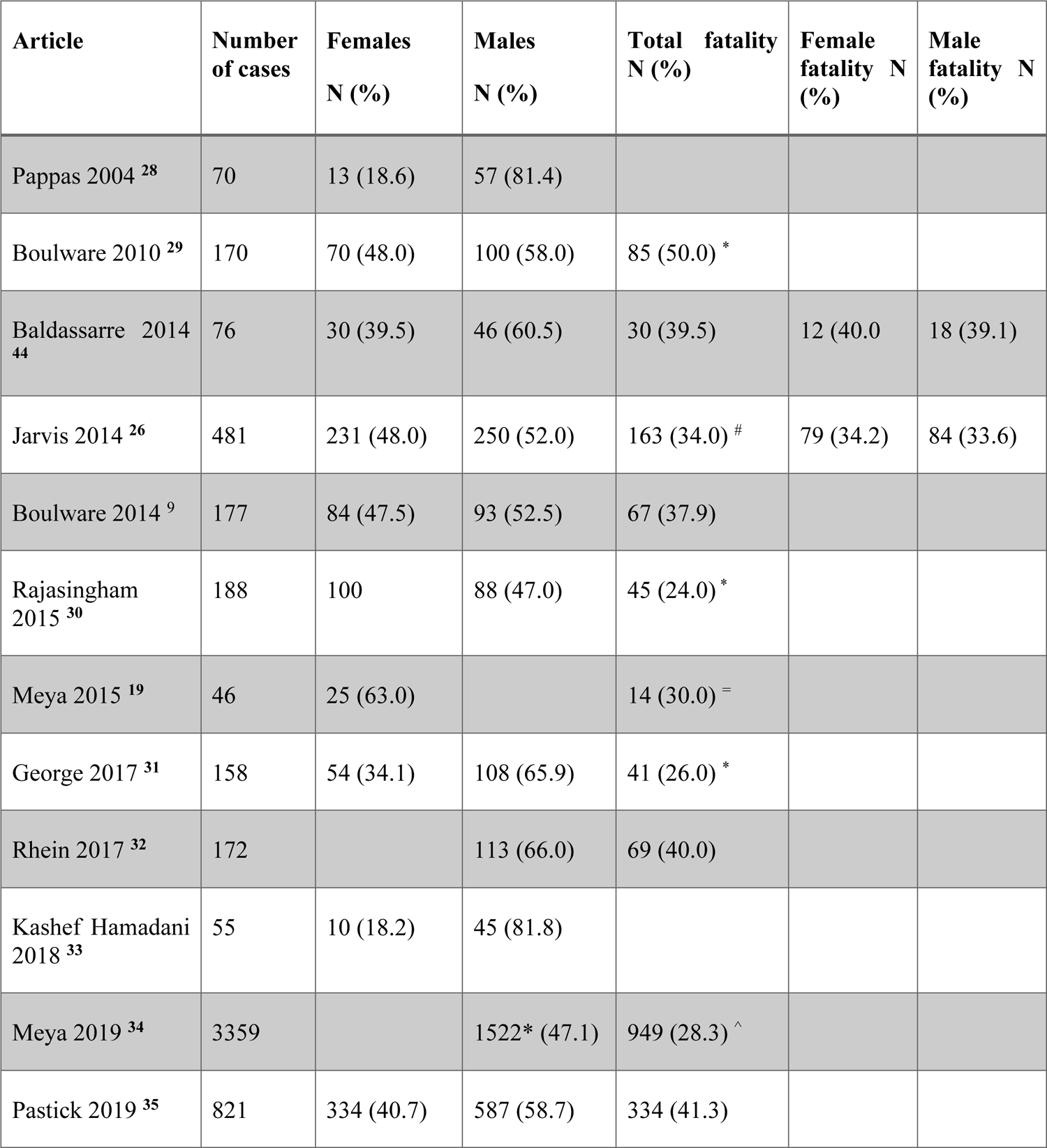
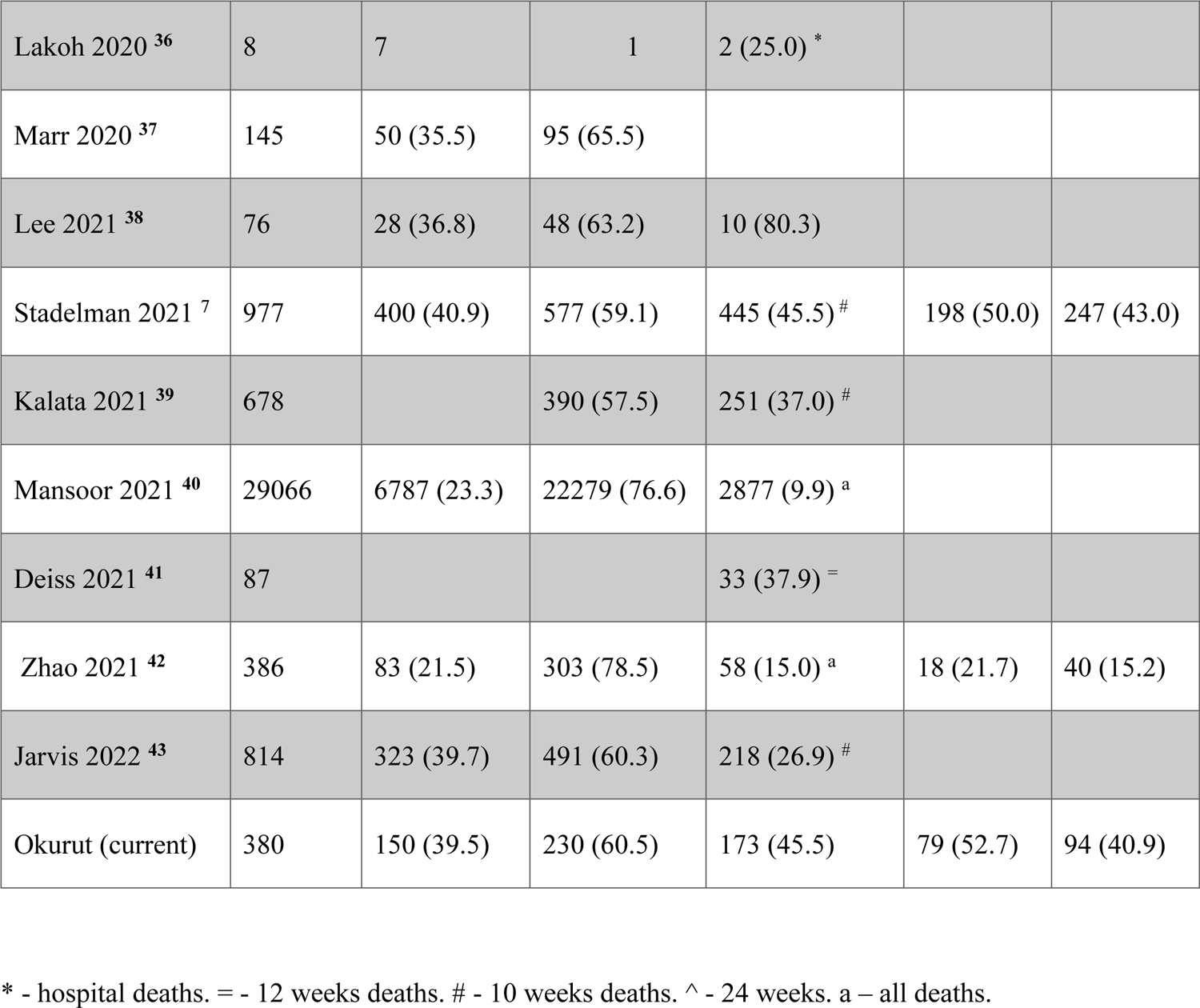
Reported gender-Associated Survival Among People with HIV-Associated Cryptococcal Meningitis.

| Article | Number of cases | Females N (%) | Males N (%) | Total fatality N (%) | Female fatality N (%) | Male fatality N (%) |
| --- | --- | --- | --- | --- | --- | --- |
| Pappas 2004 <sup>28</sup> | 70 | 13 (18.6) | 57 (81.4) |  |  |  |
| Boulware 2010 <sup>29</sup> | 170 | 70 (48.0) | 100 (58.0) | 85 (50.0) * |  |  |
| Baldassarre 2014 <sup>44</sup> | 76 | 30 (39.5) | 46 (60.5) | 30 (39.5) | 12 (40.0) | 18 (39.1) |
| Jarvis 2014 <sup>26</sup> | 481 | 231 (48.0) | 250 (52.0) | 163 (34.0) # | 79 (34.2) | 84 (33.6) |
| Boulware 2014 <sup>9</sup> | 177 | 84 (47.5) | 93 (52.5) | 67 (37.9) |  |  |
| Rajasingham 2015 <sup>30</sup> | 188 | 100 | 88 (47.0) | 45 (24.0) * |  |  |
| Meya 2015 <sup>19</sup> | 46 | 25 (63.0) |  | 14 (30.0) = |  |  |
| George 2017 <sup>31</sup> | 158 | 54 (34.1) | 108 (65.9) | 41 (26.0) * |  |  |
| Rhein 2017 <sup>32</sup> | 172 |  | 113 (66.0) | 69 (40.0) |  |  |
| Kashef Hamadani 2018 <sup>33</sup> | 55 | 10 (18.2) | 45 (81.8) |  |  |  |
| Meya 2019 <sup>34</sup> | 3359 |  | 1522* (47.1) | 949 (28.3) ^ |  |  |
| Pastick 2019 <sup>35</sup> | 821 | 334 (40.7) | 587 (58.7) | 334 (41.3) |  |  |
| Lakoh 2020 <sup>36</sup> | 8 | 7 | 1 | 2 (25.0) * |  |  |
| Marr 2020 <sup>37</sup> | 145 | 50 (35.5) | 95 (65.5) |  |  |  |
| Lee 2021 <sup>38</sup> | 76 | 28 (36.8) | 48 (63.2) | 10 (80.3) |  |  |
| Stadelman 2021 <sup>7</sup> | 977 | 400 (40.9) | 577 (59.1) | 445 (45.5) # | 198 (50.0) | 247 (43.0) |
| Kalata 2021 <sup>39</sup> | 678 |  | 390 (57.5) | 251 (37.0) # |  |  |
| Mansoor 2021 <sup>40</sup> | 29066 | 6787 (23.3) | 22279 (76.6) | 2877 (9.9) <sup>a</sup> |  |  |
| Deiss 2021 <sup>41</sup> | 87 |  |  | 33 (37.9) = |  |  |
| Zhao 2021 <sup>42</sup> | 386 | 83 (21.5) | 303 (78.5) | 58 (15.0) <sup>a</sup> | 18 (21.7) | 40 (15.2) |
| Jarvis 2022 <sup>43</sup> | 814 | 323 (39.7) | 491 (60.3) | 218 (26.9) # |  |  |
| Okurut (current) | 380 | 150 (39.5) | 230 (60.5) | 173 (45.5) | 79 (52.7) | 94 (40.9) |

\* - hospital deaths. = - 12 weeks deaths. # - 10 weeks deaths. ^ - 24 weeks. a – all deaths.

## REFERENCES

1. Wake RM, Govender NP, Omar T, et al. Cryptococcal-related mortality despite fluconazole preemptive treatment in a cryptococcal antigen screen-and-treat program. Clinical Infectious Diseases. 2020;70(8):1683–1690. doi:10.1093/cid/ciz485

2. Rhein J, Huppler Hullsiek K, Tugume L, et al. Adjunctive sertraline for HIV-associated cryptococcal meningitis: a randomised, placebo-controlled, double-blind phase 3 trial. The Lancet Infectious diseases. 2019;19(8):843–851. doi:10.1016/S1473-3099(19)30127-6

3. Hakyemez IN, Erdem H, Beraud G, et al. Prediction of unfavorable outcomes in cryptococcal meningitis: results of the multicenter Infectious Diseases International Research Initiative (ID-IRI) cryptococcal meningitis study. European Journal of Clinical Microbiology and Infectious Diseases. 2018;37(7):1231–1240. doi:10.1007/s10096-017-3142-1

4. Tenforde MW, Jarvis JN. HIV-associated cryptococcal meningitis: ongoing challenges and new opportunities. The Lancet Infectious Diseases. 2019;19(8):793–794. doi:10.1016/S1473-3099(19)30295-6

5. Stott KE, Loyse A, Jarvis JN, et al. Cryptococcal meningoencephalitis: time for action. The Lancet Infectious Diseases. 2021;21(9):e259–e271. doi:10.1016/S1473-3099(20)30771-4

6. Boulware DR, von Hohenberg M, Rolfes MA, et al. Human immune response varies by the degree of relative cryptococcal antigen shedding. Open Forum Infectious Diseases. 2016;3(1):1–7. doi:10.1093/ofid/ofv194

7. Stadelman AM, Ssebambulidde K, Tugume L, et al. Impact of biological sex on cryptococcal meningitis mortality in Uganda and South Africa. Medical Mycology. 2021;59(7):712–719. doi:10.1093/mmy/myaa108

8. Rajasingham R, Smith RM, Park BJ, et al. Global burden of disease of HIV-associated cryptococcal meningitis: an updated analysis. The Lancet Infectious diseases. 2017;17(8):873–881. doi:10.1016/S1473-3099(17)30243-8

9. Boulware DR, Meya DB, Muzoora C, et al. Timing of Antiretroviral Therapy after Diagnosis of Cryptococcal Meningitis. New England Journal of Medicine. 2014;370(26):2487–2498. doi:10.1056/nejmoa1312884

10. Kambugu A, Meya DB, Rhein J, et al. Outcomes of cryptococcal meningitis in Uganda before and after the availability of highly active antiretroviral therapy. Clinical Infectious Diseases. 2008;46(11):1694–1701. doi:10.1086/587667

11. Casadevall A, Pirofski Lanne. The damage-response framework of microbial pathogenesis. Nature reviews Microbiology. 2003;1(1):17–24. doi:10.1038/nrmicro732

12. Casadevall A, Pirofski LA. Host-pathogen interactions: Basic concepts of microbial commensalism, colonization, infection, and disease. Infection and Immunity. 2000;68(12):6511–6518. doi:10.1128/IAI.68.12.6511-6518.2000

13. Nielsen K, Cox GM, Litvintseva AP, et al. Cryptococcus neoformans α strains preferentially disseminate to the central nervous system during coinfection. Infection and Immunity. 2005;73(8):4922–4933. doi:10.1128/IAI.73.8.4922-4933.2005

14. Santiago-Tirado FH, Onken MD, Cooper JA, Klein RS, Doering TL. Trojan horse transit contributes to blood-brain barrier crossing of a eukaryotic pathogen. mBio. 2017;8(1). doi:10.1128/mBio.02183-16

15. Okurut S, Boulware DR, Olobo J, Meya DB. Landmark clinical observations and immunopathogenesis pathways linked to HIV and Cryptococcus fatal central nervous system co-infection. Mycoses. 2020;63(8):840–853. doi:10.1111/myc.13122

16. Elsegeiny W, Marr KA, Williamson PR. Immunology of Cryptococcal Infections: Developing a Rational Approach to Patient Therapy. Frontiers in immunology. 2018;9:651. doi:10.3389/fimmu.2018.00651

17. Chang CC, Omarjee S, Lim A, et al. Chemokine levels and chemokine receptor expression in the blood and the cerebrospinal fluid of HIV-infected patientswith cryptococcal meningitis and cryptococcosis-Associated immune reconstitution inflammatory syndrome. Journal of Infectious Diseases. 2013;208(10):1604–1612. doi:10.1093/infdis/jit388

18. Perera PY, Lichy JH, Waldmann TA, Perera LP. The role of interleukin-15 in inflammation and immune responses to infection: Implications for its therapeutic use. Microbes and Infection. 2012;14(3):247–261. doi:10.1016/j.micinf.2011.10.006

19. Meya DB, Okurut S, Zziwa G, et al. Cellular immune activation in cerebrospinal fluid from ugandans with cryptococcal meningitis and immune reconstitution inflammatory syndrome. Journal of Infectious Diseases. 2015;211(10):1597–1606. doi:10.1093/infdis/jiu664

20. Okurut S, Meya DB, Bwanga F, et al. B cell Compartmentalization in Blood and Cerebrospinal Fluid of HIV-Infected Ugandans with Cryptococcal Meningitis. Noverr MC, ed. Infection and Immunity. 2020;88(3):1–14. doi:10.1128/IAI.00779-19

21. Esher SK, Zaragoza O, Alspaugh JA. Cryptococcal pathogenic mechanisms: A dangerous trip from the environment to the brain. Memorias do Instituto Oswaldo Cruz. 2018;113(7):1–15. doi:10.1590/0074-02760180057

22. Mora DJ, Fortunato LR, Andrade-Silva LE, et al. Cytokine profiles at admission can be related to outcome in AIDS patients with cryptococcal meningitis. PLoS ONE. 2015;10(3):1–17. doi:10.1371/journal.pone.0120297

23. Xu XG, Pan WH, Bi XL, et al. Comparison of clinical features in patients with persistent and nonpersistent cryptococcal meningitis: Twelve years of clinical experience in four centers in china. CNS Neuroscience and Therapeutics. 2013;19(8):625–631. doi:10.1111/cns.12135

24. Rhein J, Hullsiek KH, Tugume L, et al. HHS Public Access. 2020;19(8):843–851. doi:10.1016/S1473-3099(19)30127-6.Adjunctive

25. Rolfes MA, Hullsiek KH, Rhein J, et al. The effect of therapeutic lumbar punctures on acute mortality from cryptococcal meningitis. Clinical Infectious Diseases. 2014;59(11):1607–1614. doi:10.1093/cid/ciu596

26. Jarvis JN, Bicanic T, Loyse A, et al. Determinants of mortality in a combined cohort of 501 patients with HIV-associated cryptococcal meningitis: Implications for improving outcomes. Clinical Infectious Diseases. 2014;58(5):736–745. doi:10.1093/cid/cit794

27. Lawrence DS, Leeme T, Mosepele M, Harrison TS, Seeley J, Jarvis JN. Equity in clinical trials for hiv-associated cryptococcal meningitis: A systematic review of global representation and inclusion of patients and researchers. PLoS Neglected Tropical Diseases. 2021;15(5):1–19. doi:10.1371/journal.pntd.0009376

28. Pappas PG, Bustamante B, Ticona E, et al. Recombinant Interferon-γ1b as Adjunctive Therapy for AIDS-Related Acute Cryptococcal Meningitis. The Journal of Infectious Diseases. 2004;189(12):2185–2191. doi:10.1086/420829

29. Boulware DR, Bonham SC, Meya DB, et al. Paucity of initial cerebrospinal fluid inflammation in cryptococcal meningitis is associated with subsequent immune reconstitution inflammatory syndrome. The Journal of infectious diseases. 2010;202(6):962–970. doi:10.1086/655785

30. Rajasingham R, Rhein J, Klammer K, et al. Epidemiology of meningitis in an HIV-infected Ugandan cohort. American Journal of Tropical Medicine and Hygiene. 2015;92(2):274–279. doi:10.4269/ajtmh.14-0452

31. George IA, Santos CAQQ, Olsen MA, Powderly WG. Epidemiology of cryptococcosis and cryptococcal meningitis in a large retrospective cohort of patients after solid organ transplantation. Open Forum Infectious Diseases. 2017;4(1):1–7. doi:10.1093/ofid/ofx004

32. Rhein J, Morawski BM, Hullsiek KH, et al. Associated Cryptococcal Meningitis: an open-label dose-ranging study. Lancet Infect Dis. 2017;16(7):809–818. doi:10.1016/S1473-3099(16)00074-8.Efficacy

33. Kashef Hamadani BH, Franco-Paredes C, McCollister B, Shapiro L, Beckham JD, Henao-Martínez AF. Cryptococcosis and cryptococcal meningitis: New predictors and clinical outcomes at a United States academic medical centre. Mycoses. 2018;61(5):314–320. doi:10.1111/myc.12742

34. Meya DB, Kiragga AN, Nalintya E, … Reflexive laboratory-based cryptococcal antigen screening and preemptive fluconazole therapy for cryptococcal antigenemia in HIV-infected individuals with …. Journal of acquired …. 2019;80(2):182–189.

35. Pastick KA, Bangdiwala AS, Abassi M, et al. Seizures in Human Immunodeficiency Virus-Associated Cryptococcal Meningitis: Predictors and Outcomes. Open Forum Infectious Diseases. 2019;6(11):1–7. doi:10.1093/ofid/ofz478

36. Lakoh S, Rickman H, Sesay M, et al. Prevalence and mortality of cryptococcal disease in adults with advanced HIV in an urban tertiary hospital in Sierra Leone: A prospective study. BMC Infectious Diseases. 2020;20(1):1–7. doi:10.1186/s12879-020-4862-x

37. Marr KA, Sun Y, Spec A, et al. A multicenter, longitudinal cohort study of cryptococcosis in human immunodeficiency virus-negative people in the United States. Clinical Infectious Diseases. 2020;70(2):252–261. doi:10.1093/cid/ciz193

38. Lee WJ, Ryu YJ, Moon J, et al. Enlarged periventricular space and periventricular lesion extension on baseline brain MRI predicts poor neurological outcomes in cryptococcus meningoencephalitis. Scientific Reports. 2021;11(1):1–11. doi:10.1038/s41598-021-85998-6

39. Kalata N, Ellis J, Kanyama C, et al. Short-term Mortality outcomes of HIV-Associated cryptococcal meningitis in antiretroviral therapy-naïve and -experienced patients in sub-saharan Africa. Open Forum Infectious Diseases. 2021;8(10):1–5. doi:10.1093/ofid/ofab397

40. Mansoor A e.Rehman, Thompson J, Sarwari AR. Delays in lumbar puncture are independently associated with mortality in cryptococcal meningitis: a nationwide study. Infectious Diseases. 2021;53(5):361–369. doi:10.1080/23744235.2021.1889656

41. Deiss R, Loreti CV, Gutierrez AG, et al. High burden of cryptococcal antigenemia and meningitis among patients presenting at an emergency department in Maputo, Mozambique. PLoS ONE. 2021;16(4 April):1-11. doi:10.1371/journal.pone.0250195

42. Zhao T, Xu XL, Nie JM, et al. Establishment of a novel scoring model for mortality risk prediction in HIV-infected patients with cryptococcal meningitis. BMC Infectious Diseases. 2021;21(1):1–10. doi:10.1186/s12879-021-06417-9

43. Jarvis JN, Lawrence DS, Meya DB, et al. Single-Dose Liposomal Amphotericin B Treatment for Cryptococcal Meningitis. New England Journal of Medicine. 2022;386(12):1109–1120. doi:10.1056/nejmoa2111904

44. Baldassarre R, Mdodo R, Omonge E, et al. Mortality after clinical management of aids-associated cryptococcal meningitis in Kenya. East African Medical Journal. 2014;91(5):145–151.

45. Furman D, Hejblum BP, Simon N, et al. Systems analysis of sex differences reveals an immunosuppressive role for testosterone in the response to influenza vaccination. Proceedings of the National Academy of Sciences of the United States of America. 2014;111(2):869–874. doi:10.1073/pnas.1321060111

46. Fink AL, Engle K, Ursin RL, Tang WY, Klein SL. Biological sex affects vaccine efficacy and protection against influenza in mice. Proceedings of the National Academy of Sciences of the United States of America. 2018;115(49):12477–12482. doi:10.1073/pnas.1805268115

47. Garelnabi M, Taylor-Smith LM, Bielska E, Hall RA, Stones D, May RC. Quantifying donor-to-donor variation in macrophage responses to the human fungal pathogen Cryptococcus neoformans. PLoS ONE. 2018;13(3):1–12. doi:10.1371/journal.pone.0194615

48. Rhein J, Morawski BM, Hullsiek KH, et al. Efficacy of adjunctive sertraline for the treatment of HIV-associated cryptococcal meningitis: an open-label dose-ranging study. The Lancet Infectious diseases. 2016;16(7):809–818. doi:10.1016/S1473-3099(16)00074-8

49. Vidal JE, Toniolo C, Paulino A, et al. Performance of cryptococcal antigen lateral flow assay in serum, cerebrospinal fluid, whole blood, and urine in HIV-infected patients with culture-proven cryptococcal meningitis admitted at a Brazilian referral center. Revista do Instituto de Medicina Tropical de Sao Paulo. 2018;60:e1–e1. doi:10.1590/s1678-9946201860001

50. Rajasingham R, Wake RM, Beyene T, Katende A, Letang E, Boulware DR. Cryptococcal Meningitis Diagnostics and Screening in the Era of Point-of-Care Laboratory Testing. Journal of clinical microbiology. 2019;57(1):e01238–18. doi:10.1128/JCM.01238-18

51. Vazirinejad R, Ahmadi Z, Arababadi MK, Hassanshahi G, Kennedy D. The biological functions, structure and sources of CXCL10 and its outstanding part in the pathophysiology of multiple sclerosis. NeuroImmunoModulation. 2014;21(6):322–330. doi:10.1159/000357780

52. Michlmayr D, McKimmie CS. Role of CXCL10 in central nervous system inflammation. International Journal of Interferon, Cytokine and Mediator Research. 2014;6(1):1–18. doi:10.2147/IJICMR.S35953

53. Teixeira AL, Gama CS, Rocha NP, Teixeira MM. Revisiting the role of eotaxin-1/CCL11 in psychiatric disorders. Frontiers in Psychiatry. 2018;9(JUN):1–6. doi:10.3389/fpsyt.2018.00241

54. Parajuli B, Horiuchi H, Mizuno T, Takeuchi H, Suzumura A. CCL11 enhances excitotoxic neuronal death by producing reactive oxygen species in microglia. Glia. 2015;63(12):2274–2284. doi:10.1002/glia.22892

55. Mamik MK, Ghorpade A. Chemokine CXCL8 promotes HIV-1 replication in human monocyte-derived macrophages and primary microglia via nuclear factor-κB pathway. PLoS ONE. 2014;9(3):1–13. doi:10.1371/journal.pone.0092145

56. Chauhan P, Lokensgard JR. Glial cell expression of PD-L1. International Journal of Molecular Sciences. 2019;20(7):1–12. doi:10.3390/ijms20071677

57. Burmeister AR, Marriott I. The interleukin-10 family of cytokines and their role in the CNS. Frontiers in Cellular Neuroscience. 2018;12(November):1–13. doi:10.3389/fncel.2018.00458

58. Ringnér M. What is principal component analysis? Nature Biotechnology. 2008;26(3):303–304. doi:10.1038/nbt0308-303

59. Giuliani A. The application of principal component analysis to drug discovery and biomedical data. Drug Discovery Today. 2017;22(7):1069–1076. doi:10.1016/j.drudis.2017.01.005

60. Scriven JE, Graham LM, Schutz C, et al. A glucuronoxylomannan-associated immune signature, characterized by monocyte deactivation and an increased interleukin 10 level, is a predictor of death in cryptococcal meningitis. Journal of Infectious Diseases. 2016;213(11):1725–1734. doi:10.1093/infdis/jiw007

61. Gerstein AC, Jackson KM, McDonald TR, et al. Identification of pathogen genomic differences that impact human immune response and disease during cryptococcus neoformans infection. mBio. 2019;10(4):1–22. doi:10.1128/mBio.01440-19

62. Traino K, Snow J, Ham L, et al. HIV-Negative Cryptococcal Meningoencephalitis Results in a Persistent Frontal-Subcortical Syndrome. Scientific Reports. 2019;9(1):1–9. doi:10.1038/s41598-019-54876-7

63. Vanherp L, Poelmans J, Weerasekera A, et al. Trehalose as quantitative biomarker for in vivo diagnosis and treatment follow-up in cryptococcomas. Translational research: the journal of laboratory and clinical medicine. 2021;230:111–122. doi:10.1016/j.trsl.2020.11.001

64. Leite AGB, Vidal JE, Bonasser Filho F, Nogueira RS, de Oliveira ACP. Cerebral infarction related to cryptococcal meningitis in an HIV-infected patient: case report and literature review. The Brazilian journal of infectious diseases: an official publication of the Brazilian Society of Infectious Diseases. 2004;8(2):175–179. doi:10.1590/S1413-86702004000200008

65. Orsini J, Blaak C, Mahmoud D, Young-Gwang J. Massive cerebral edema resulting in brain death as a complication of cryptococcus neoformans meningitis. Journal of Community Hospital Internal Medicine Perspectives. 2015;5(1):3–6. doi:10.3402/jchimp.v5.26098

66. Jarvis JN, Meintjes G, Bicanic T, et al. Cerebrospinal Fluid Cytokine Profiles Predict Risk of Early Mortality and Immune Reconstitution Inflammatory Syndrome in HIV-Associated Cryptococcal Meningitis. PLoS Pathogens. 2015;11(4):1–17. doi:10.1371/journal.ppat.1004754

67. Jarvis JN, Casazza JP, Stone HH, et al. The phenotype of the cryptococcus-specific CD4+ memory T-cell response is associated with disease severity and outcome in HIV-associated cryptococcal meningitis. Journal of Infectious Diseases. 2013;207(12):1817–1828. doi:10.1093/infdis/jit099

68. Qiao X, Zhang W, Zhao W. Role of CXCL10 in Spinal Cord Injury. Int J Med Sci. 2022;19(14):2058–2070. doi:10.7150/ijms.76694

69. Teixeira AL, Gama CS, Rocha NP, Teixeira MM. Revisiting the Role of Eotaxin-1/CCL11 in Psychiatric Disorders. Front Psychiatry. 2018;9:241. doi:10.3389/fpsyt.2018.00241

70. Okafor EC, Hullsiek KH, Williams DA, et al. Correlation between blood and CSF compartment cytokines and chemokines in subjects with cryptococcal meningitis. Mediators of Inflammation. 2020;2020. doi:10.1155/2020/8818044

71. Boulware DR, Meya DB, Bergemann TL, et al. Clinical features and serum biomarkers in HIV immune reconstitution inflammatory syndrome after cryptococcal meningitis: A prospective cohort study. PLoS Medicine. 2010;7(12):1–14. doi:10.1371/journal.pmed.1000384

72. Lobo-Silva D, Carriche GM, Castro AG, Roque S, Saraiva M. Balancing the immune response in the brain: IL-10 and its regulation. Journal of Neuroinflammation. 2016;13(1):1–10. doi:10.1186/s12974-016-0763-8

73. Wake RM, Govender NP, Omar T, et al. Cryptococcal-related mortality despite fluconazole pre-emptive treatment in a cryptococcal antigen (CrAg) screen-and-treat programme. Clinical infectious diseases: an official publication of the Infectious Diseases Society of America. Published online June 2019. doi:10.1093/cid/ciz485

74. Meya D, Okurut S, Zziwa G, et al. Monocyte Phenotype and IFN-γ-Inducible Cytokine Responses Are Associated with Cryptococcal Immune Reconstitution Inflammatory Syndrome. Journal of Fungi. 2017;3(2):28. doi:10.3390/jof3020028

75. Meya DB, Okurut S, Zziwa G, Cose S, Boulware DR, Janoff EN. HIV-Associated Cryptococcal Immune Reconstitution Inflammatory Syndrome Is Associated with Aberrant T Cell Function and Increased Cytokine Responses. *Journal of fungi (Basel*, Switzerland). 2019;5(2):42. doi:10.3390/jof5020042

76. Bickel M. The role of interleukin-8 in inflammation and mechanisms of regulation. Journal of periodontology. 1993;64(5 Suppl):456–460.

77. Ehrlich LC, Hu S, Sheng WS, et al. Cytokine regulation of human microglial cell IL-8 production. Journal of immunology (Baltimore, Md: 1950). 1998;160(4):1944–1948.

78. Rosales C. Neutrophil: A cell with many roles in inflammation or several cell types? Frontiers in Physiology. 2018;9(FEB):1–17. doi:10.3389/fphys.2018.00113

79. Guimarães-Costa AB, Nascimento MTC, Wardini AB, Pinto-Da-Silva LH, Saraiva EM. ETosis: A microbicidal mechanism beyond cell death. Journal of Parasitology Research. 2012;2012(Table 1). doi:10.1155/2012/929743

80. Musubire AK, Meya DB, Rhein J, et al. Blood neutrophil counts in HIV-infected patients with cryptococcal meningitis: Association with mortality. PloS one. 2018;13(12):e0209337–e0209337. doi:10.1371/journal.pone.0209337

81. Grahl N, Shepardson KM, Chung D, Cramer RA. Hypoxia and fungal pathogenesis: To air or not to air? Eukaryotic Cell. 2012;11(5):560–570. doi:10.1128/EC.00031-12

82. Diehl JW, Hullsiek KH, Okirwoth M, et al. Cerebral Oximetry for Detecting High-mortality Risk Patients with Cryptococcal Meningitis. Published online 2017:1–7. doi:10.1093/ofid/ofy105

83. Shimoda Y, Ohtomo S, Arai H, Ohtoh T, Tominaga T. Subarachnoid small vein occlusion due to inflammatory fibrosis—a possible mechanism for cerebellar infarction in cryptococcal meningoencephalitis: a case report. BMC Neurology. 2017;17(1):157. doi:10.1186/s12883-017-0934-y

84. Hung CW, Chang WN, Kung CT, et al. Predictors and long-term outcome of seizures in human immuno-deficiency virus (HIV)-negative cryptococcal meningitis. BMC Neurology. 2014;14(1):1–8. doi:10.1186/s12883-014-0208-x

85. Saito T, Saido TC. Neuroinflammation in mouse models of Alzheimer’s disease. Clinical and Experimental Neuroimmunology. 2018;9(4):211–218. doi:10.1111/cen3.12475

86. Lu Y, Ma C, Chen R, et al. Development and validation of a new scoring system for the early diagnosis of tuberculous meningitis in adults. Diagnostic Microbiology and Infectious Disease. 2021;101(2):115393. doi:10.1016/j.diagmicrobio.2021.115393

87. D’Inzeo T, Menchinelli G, De Angelis G, et al. Implementation of the eazyplex® CSF direct panel assay for rapid laboratory diagnosis of bacterial meningitis: 32-month experience at a tertiary care university hospital. European Journal of Clinical Microbiology and Infectious Diseases. 2020;39(10):1845–1853. doi:10.1007/s10096-020-03909-5

88. Sokol CL, Luster AD. The chemokine system in innate immunity. Cold Spring Harbor Perspectives in Biology. 2015;7(5):1–20. doi:10.1101/cshperspect.a016303

89. Veenstra M, León-Rivera R, Li M, Gama L, Clements JE, Berman JW. Mechanisms of CNS viral seeding by HIV+ CD14+ CD16+ monocytes: Establishment and reseeding of viral reservoirs contributing to HIV-associated neurocognitive disorders. mBio. 2017;8(5):1–15. doi:10.1128/mBio.01280-17

90. Persidsky Y, Poluektova L. Immune Privilege and HIV-1 Persistence in the CNS. Vol 213.; 2006:180–194. doi:10.1111/j.1600-065X.2006.00440.x

91. Panackal AA, Wuest SC, Lin YC, et al. Paradoxical Immune Responses in Non-HIV Cryptococcal Meningitis. PLoS Pathogens. 2015;11(5):1–27. doi:10.1371/journal.ppat.1004884

92. Kawakami K, Qureshi MH, Koguchi Y, Nakajima K, Saito A. Differential effect of Cryptococcus neoformans on the production of IL-12p40 and IL-10 by murine macrophages stimulated with lipopolysaccharide and gamma interferon. FEMS Microbiology Letters. 1999;175(1):87–94. doi:10.1016/S0378-1097(99)00172-X

93. Trinchieri G. Interleukin-12 and the regulation of innate resistance and adaptive immunity. Nature Reviews Immunology. 2003;3(2):133–146. doi:10.1038/nri1001

94. Kuwabara T, Ishikawa F, Kondo M, Kakiuchi T. The Role of IL-17 and Related Cytokines in Inflammatory Autoimmune Diseases. Mediators of Inflammation. 2017;2017. doi:10.1155/2017/3908061

95. Lee SC, Casadevall A, Dickson DW. Immunohistochemical localization of capsular polysaccharide antigen in the central nervous system cells in cryptococcal meningoencephalitis. The American journal of pathology. 1996;148(4):1267–1274.

96. Okurut S, Okafor E, Boulware DR, et al. Okurut, S., Okafor, E., Boulware, RD., Rhein J., Manabe, CY., Olobo OJ., Meya BD., and Janoff, NE., (2023). Divergent neuroimmune signatures and HIV-associated cryptococcal meningitis survival. Abstract and poster retrieved on July 28, 2023 from CROI webseite. 489_ePoster. 2023;(2023). https://www.croiconference.org/abstract/divergent-neuroimmune-signatures-and-hiv-associated-cryptococcal-meningitis-survival/

97. Okurut, S. (2023) B cell responses, immune modulation and survival among patients with HIV-associated cryptococcal meningitis. [PhD. Thesis]. [Kampala, Uganda]: Makerere University.

